# Impacts of vaccination and asymptomatic testing on SARS-CoV-2 transmission dynamics in a university setting

**DOI:** 10.1101/2021.11.22.21266565

**Authors:** Emily Nixon, Amy Thomas, Daniel Stocks, Antoine M.G. Barreaux, Gibran Hemani, Adam Trickey, Rachel Kwiatkowska, Josephine Walker, David Ellis, Leon Danon, Caroline Relton, Hannah Christensen, Ellen Brooks-Pollock

**Affiliations:** School of Biological Sciences, University of Bristol, Bristol, UK; Bristol Veterinary School, University of Bristol, Bristol, UK; Population Health Sciences, University of Bristol, Bristol, UK; MRC Integrative Epidemiology Unit, University of Bristol, Bristol, UK; NIHR Health Protection Research Unit in Behavioural Science and Evaluation at University of Bristol, Bristol, UK; School of Mathematics, University of Bristol, Bristol, UK; Department of Engineering Mathematics, University of Bristol, Bristol, UK

**Author notes:** Corresponding author Corresponding author, Corresponding author address: School of Biological Sciences, 24 Tyndall Avenue, Bristol, BS8 1TQ.

## Abstract

We investigate the impact of vaccination and asymptomatic testing uptake on SARS-CoV-2 transmission in a university student population using a stochastic compartmental model. We find that the magnitude and timing of outbreaks is highly variable depending on the transmissibility of the most dominant strain of SARS CoV-2 and under different vaccine uptake levels and efficacies. When delta is the dominant strain, low level interventions (no asymptomatic testing, 30% vaccinated with a vaccine that is 80% effective at reducing infection) lead to 53-71% of students become infected during the first term. Asymptomatic testing is most useful when vaccine uptake is low: when 30% of students are vaccinated, 90% uptake of asymptomatic testing leads to almost half the case numbers. With high interventions (90% using asymptomatic testing, 90% vaccinated) cumulative incidence is 7-9%, with around 80% of these cases estimated to be asymptomatic. However, under emergence of a new variant that is at least twice as transmissible as delta and with the vaccine efficacy against infection reduced to 55%, large outbreaks are likely in universities, even with very high (90%) uptake of vaccination and 100% uptake of asymptomatic testing. If vaccine efficacy against infection against this new variant is higher (70%), then outbreaks can be mitigated if there is least 50% uptake of asymptomatic testing additional to 90% uptake of vaccination. Our findings suggest that effective vaccination is critical for controlling SARS-CoV-2 transmission in university settings with asymptomatic testing ranging from additionally useful to critical, depending on effectiveness and uptake of vaccination. Other measures may be necessary to control outbreaks under the emergence of a more transmissible variant with vaccine escape.

## Introduction

The university setting presents a unique environment for infectious disease transmission - a primarily young demographic, with high-density living arrangements and large numbers of social contacts, as well as mass migrations of international and domestic students at the beginning and end of university terms. University student populations are interlinked with university staff, and often also with the wider community surrounding the university, which can have implications for disease transmission in the community. During the current pandemic, understanding the extent of transmission of COVID-19 in university settings has therefore been an important public health goal.

There are several key factors that influence the extent of transmission of COVID-19 in a university setting: social contact patterns, non-pharmaceutical interventions (NPIs, such as the wearing of face-coverings), mass asymptomatic testing (uptake and effectiveness), vaccine uptake and effectiveness, levels of pre-existing immunity, and the robustness of vaccine-induced immunity. During the COVID-19 pandemic there have been several interventions to control transmission at universities, including periods of suspension of in-person activities and teaching. For example, in the first UK lockdown, all students, other than those who were undertaking practical courses which require specialist equipment, had their learning delivered remotely and were able to return to a non-term residence^1^. When in-person activities and teaching resumed, NPIs, predominately social distancing and mask wearing, were recommended to limit SARS-CoV-2 transmission^2^. Despite these NPIs being in place during Autumn 2020, outbreaks of COVID-19 were observed at several UK universities, with the largest of these occurring in halls of residence^3^.

Mass testing of asymptomatic individuals using lateral flow tests (LFT) is another measure that has been used by universities to help mitigate the impact of COVID-19^4^ and in the autumn term of 2021/2022 has been recommended by the UK Department of Education for the reopening of higher education institutions^5^. Testing of asymptomatic individuals leads to increased case detection and may be particularly important in the university student population where, due to the younger age structure, cases are more likely to be asymptomatic^6,7^. Previous modelling of asymptomatic testing in a university found that adherence to testing and isolation was important for reducing transmission^8^, particularly at high values of the reproduction number^8-10^.

In the UK, a high proportion of university students are expected to have been vaccinated prior to the start of autumn term of 2021/2022 (at least 55% with two doses and at least 65% with one dose^11^), and some universities have been running vaccination programs to encourage those who have not yet been vaccinated, or who have come from overseas, to get vaccinated at the start of term^12^. However, in a university context, the impact of vaccination under different levels of uptake is unknown, the effect of mass testing programmes is under-explored and the combination of the two interventions in parallel may have uncharacterised effects. One agent-based model based on a US campus has looked at a range of vaccination scenarios with different efficacies, alone and in combination with other mitigation strategies: however, the study does not consider the impact of varying vaccine uptake levels^13^. There have also been no investigations in a university setting of the impact of variants such as the omicron (B.1.1529) variant which could be more transmissible than the delta variant (B.1.617.2) and may reduce vaccine efficacy against infection.

Immune responses to SARS-CoV-2 following infection or vaccination have been shown to wane. Accurate estimates for rates of waning immunity are challenging due to heterogeneity in observed responses and understanding how these immune responses correlate to protection against (re)infection and disease^14^. Based on observational studies, the risk of reinfection is reduced by 80-93% for at least 6-9 months following confirmed infection^15-19^. Vaccine-induced immunity has been demonstrated to remain robust for at least 4 months following two doses of BNT162b2 mRNA COVID-19 vaccine, with assessments in vaccine efficacy against infection and severe disease showing a decline over time^20^.

To help institutions set policies to control COVID-19, we adapted an existing stochastic compartmental model of SARS-CoV-2 transmission in a university student population^9^ to investigate the impact of uptake and effectiveness of vaccination and asymptomatic testing in this setting.

## Results

Using a stochastic compartmental model of SARS-CoV-2 transmission in a university setting (Figure 1), we find low levels of vaccination uptake (30%) with a vaccine that is 80% effective at reducing infection) are insufficient to prevent outbreaks amongst students in the Autumn term of 2021 (Figure 2) where delta (B.1.617.2) is the most dominant variant in the population. With no asymptomatic testing and at this low level of vaccination uptake, the cumulative incidence for the term is estimated to be between 53-71% (14,901-19,962 students in the population modelled) depending on the number of students initially infected at the start of term and under the assumption that importation of disease from the community occurs, with a peak in community cases expected in the winter months (Figure 3). The inclusion of asymptomatic testing with low level vaccination (30%) can reduce the cumulative incidence to 37-56% (10,403-15,745 students) when 90% of students are doing a LFT twice per week, 4 days apart (Figure 3). At these low levels of vaccination uptake (30%), the outbreak peak occurs around the end of November or early December (the end of term), with the time of the peak dependent on the asymptomatic testing uptake (Figure 2). There are approximately 1000 students infected at the peak of the outbreak when there is high uptake of asymptomatic testing (90%), 1300 students when there is medium uptake (50%), 1750 students with low uptake (20%), and 2250 students when there is no asymptomatic testing (Figure 2). At the peak of the outbreak, there could be up to 1405 (5%) students isolating on any one day when there are high levels of asymptomatic testing uptake (90%). With no asymptomatic testing, only up to 352 (1.3%) students would be isolating at the peak since fewer infections would be detected (Figure 4).

**Figure 1.**
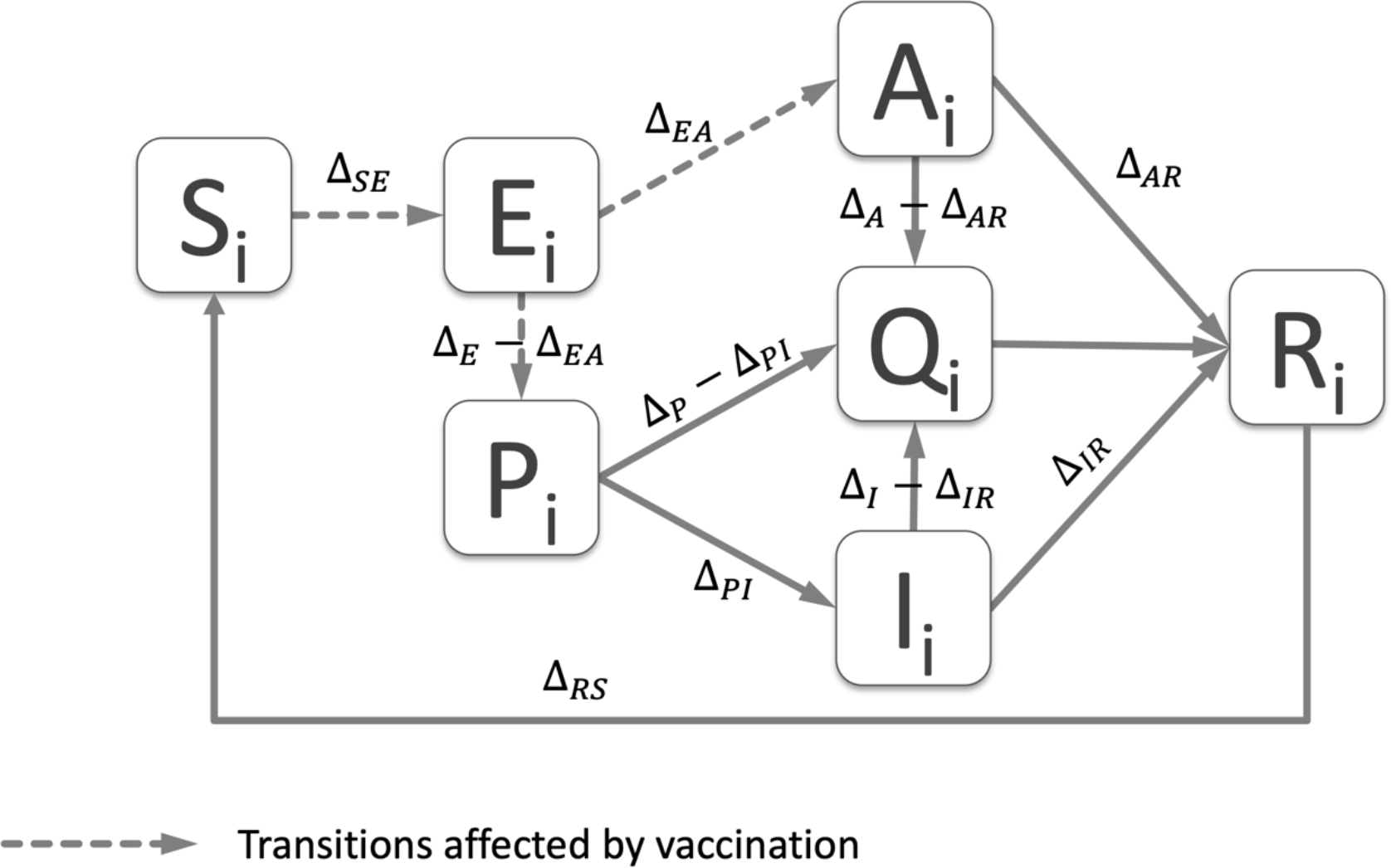
Model schematic showing the disease states and rates between them in the stochastic compartmental model. The disease states are S: susceptible to infection, E: exposed, or infected but pre-infectious, P: pre-symptomatic and infectious, I: symptomatic and infectious, A: asymptomatic and infectious, Q: in quarantine, R: recovered and immune. The subscript *i* refers to the subgroup. The model is adapted from an existing model for COVID-19 transmission in university student population, fully described in Brooks Pollock et al., 2021^9^, where the rates are explained in full and shown in equations (1) and (2) and Table 1 of that study. Here, waning immunity from natural infection is added to the model by the addition of a transition from the R to S disease state and the impact of vaccination is incorporated. The transitions which are affected by vaccination are indicated by the dotted lines and described here in the main text of the methods and in equations 1, 2 and 3.

**Figure 2.**
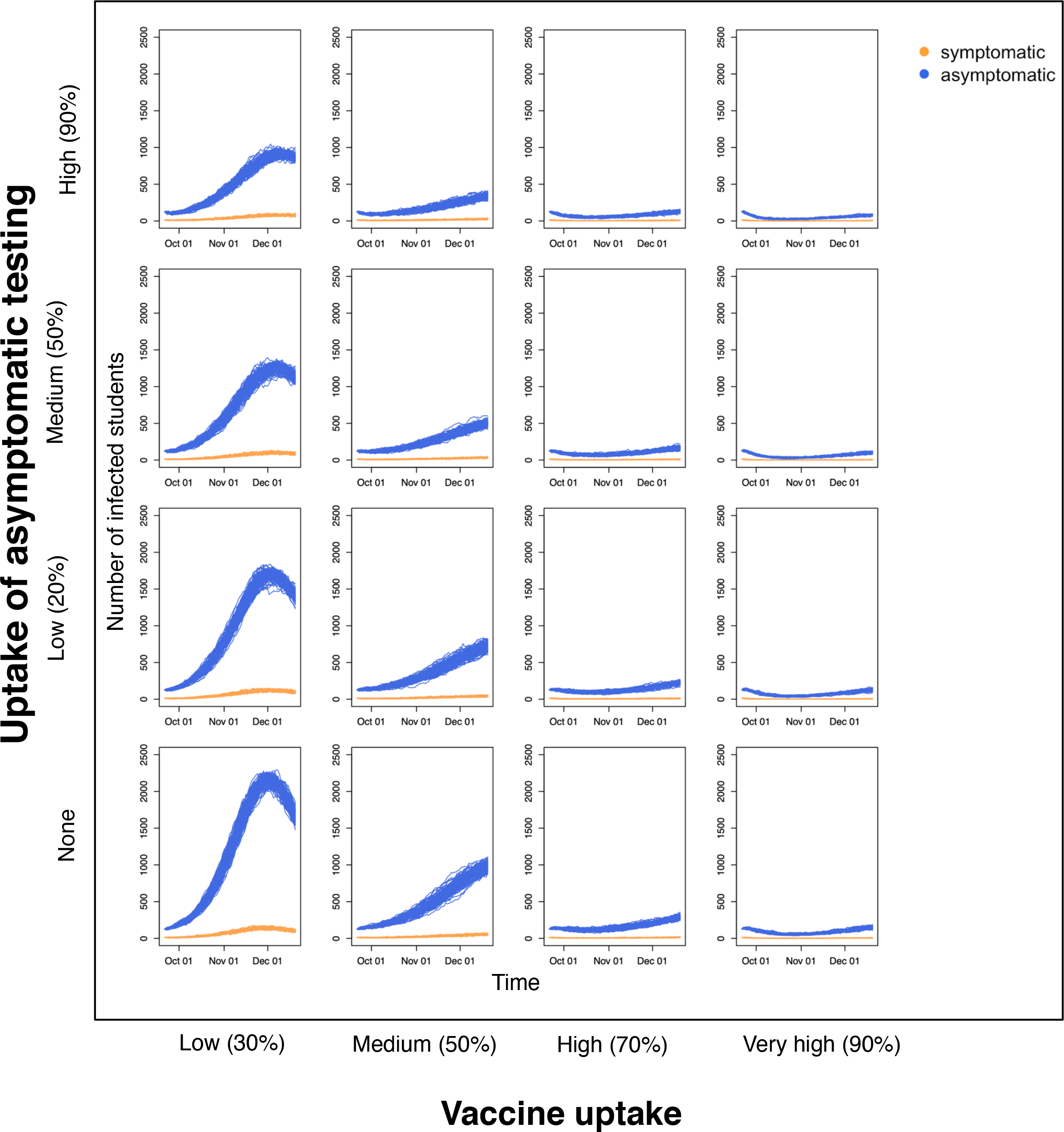
Daily numbers of symptomatic and asymptomatic SARS-CoV-2 infections amongst students under different levels of vaccine uptake and asymptomatic testing. Scenarios of varying levels of vaccine uptake and asymptomatic testing were considered in a population size of 28116 students. The daily number of students who are infected with SARS-CoV-2 in the autumn term of 2021/2022 and are symptomatic are shown in orange, asymptomatic infections are shown in blue. Importation from the community was assumed to occur at a time-varying seasonal rate with maximum value of 1e-3. Simulations were started with 1% of students initially infected and 10% having natural immunity. Results of the given scenarios are shown for 100 stochastic repeats.

**Figure 3.**
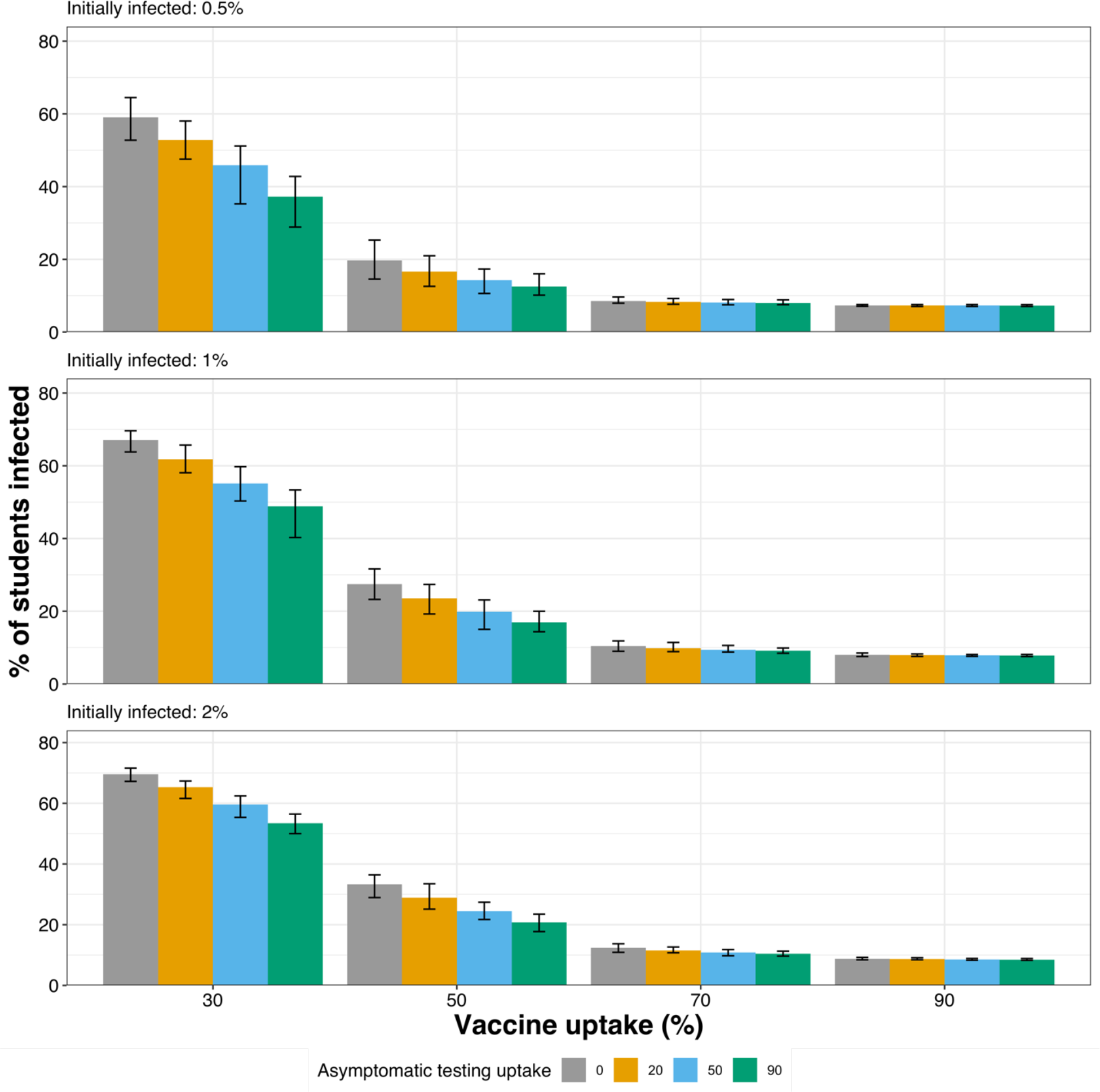
The cumulative percentage of university students infected with SARS-CoV-2 in the Autumn term of 2021 as predicted under different scenarios of asymptomatic testing uptake, vaccine uptake and percentage of initially infected students at the start of term. Each scenario is repeated 100 times, the mean and range are given. The results shown do not include those who were infected but whose immunity has waned by the end of term. Importation from the community was assumed to occur at a time-varying seasonal rate with maximum value of 1e-3.

**Figure 4.**
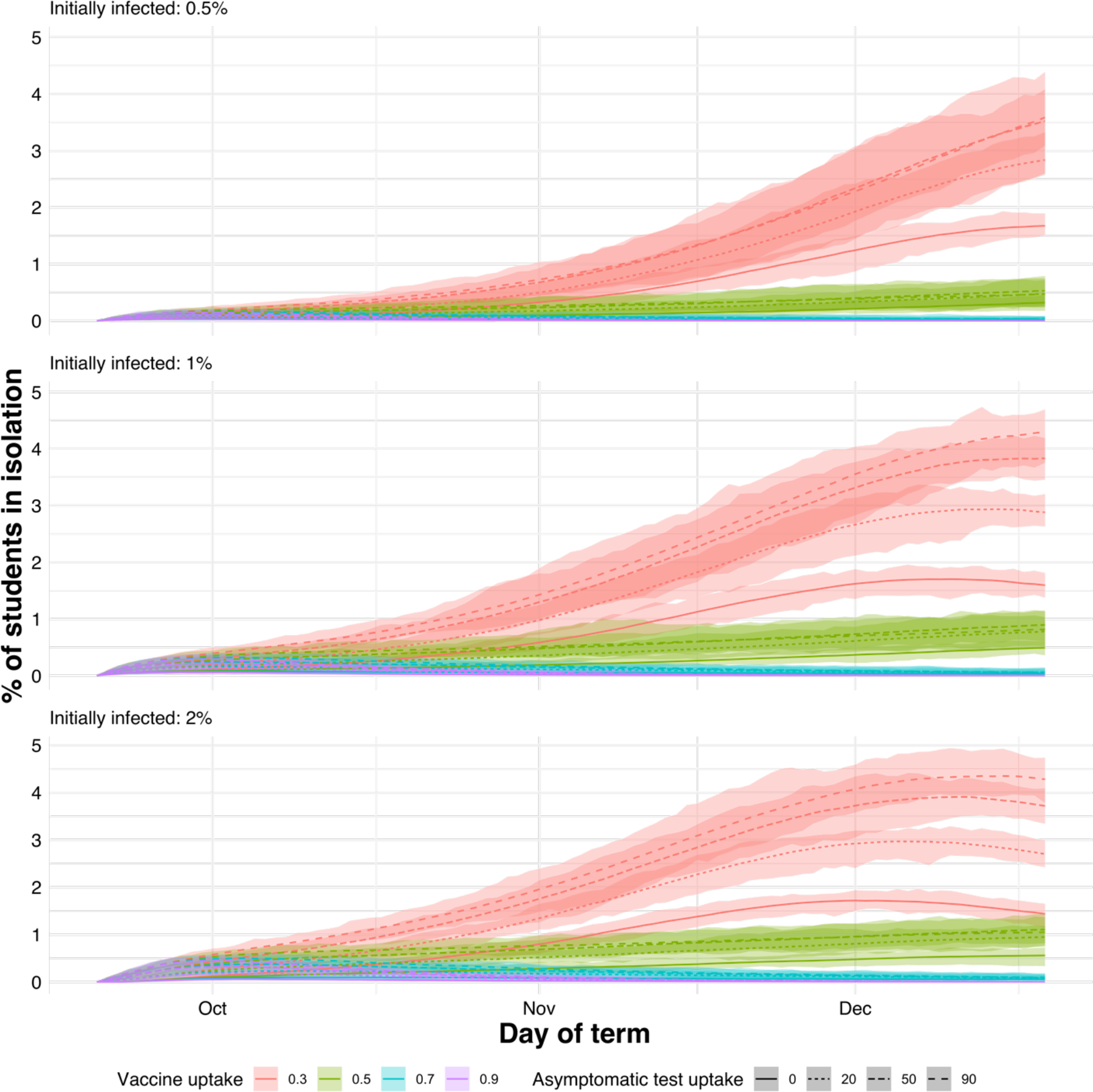
The percentage of students in a university population isolating each day after testing positive for SARS-CoV-2 in the Autumn term of 2021 as predicted under different scenarios of asymptomatic testing uptake, vaccine uptake and percentage of initially infected students at the start of term. Each scenario is repeated 100 times and the mean and the range is given.

With medium levels of vaccination uptake (50%) outbreaks still occur in the university student population (Figure 2), although they lead to approximately half the number of students infected compared to low (30%) uptake (Figure 3). The inclusion of asymptomatic testing with a medium level of vaccination uptake can reduce the cumulative incidence over the autumn term by up to 19.7% (5249 fewer students infected) when 90% of students are taking a LFT twice per week, 4 days apart (Figure 3). At medium levels of vaccination uptake, the outbreak does not reach its peak before the end of term (Figure 2) and the number of students isolating per day never exceeds 1.5% (421 students, Figure 4).

With high (70%) to very high (90%) levels of vaccination uptake, outbreaks are largely contained (Figure 2) with a range of 7.5-13.7% (2108-3851) students becoming infected in the autumn term when there is 70% uptake and 7.0-9.2% when there is 90% uptake (Figure 3). These results are based on the assumption that importation from the community occurs, and that there is an increase in community cases during the winter months which is reflected in the student population. However, when there is no importation of COVID-19 from the community into the student population and there is 90% vaccination uptake in students, there is no increase in cases in the winter months at the end of term (Figure 5).

**Figure 5.**
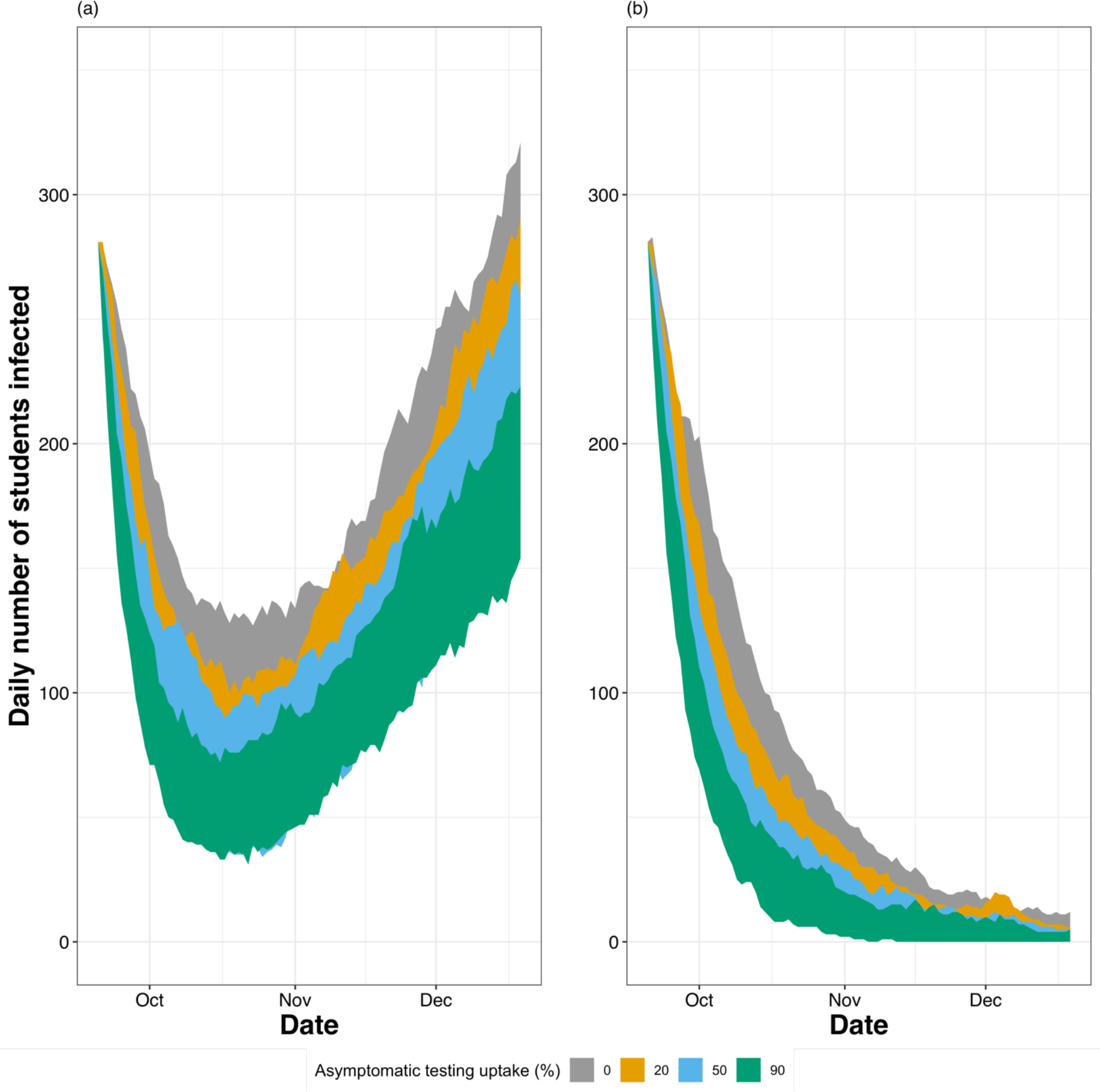
Daily numbers of SARS-CoV-2 infections in a population of students (n=28116) during the autumn term of 2021 in model simulations where 90% of students are vaccinated under different levels of asymptomatic testing and either (a) importation of infection occurs from the community or (b) no importation occurs. It is assumed those who are vaccinated have received two doses of a mRNA vaccine, with the second dose being administered at least 2 weeks prior to the start of term. Importation from the community was assumed to occur at a time-varying seasonal rate with more cases in winter and a maximum value of 1e-3. Simulations were initialised with 1% of students infected and 10% having natural immunity. Results of the given scenarios are shown for 100 stochastic repeats.

At very high (90%) levels of vaccination uptake, asymptomatic testing reduces case numbers by 2.2%, which in the student population modelled here is up to 613 students. The impact of asymptomatic testing is greater the lower the vaccination uptake, causing up to a 6.2% reduction at high levels of vaccination uptake (70%), 26.2% at medium vaccination levels (50% uptake) and a 42.7% reduction in cases at low vaccination levels (30% uptake) (Figure 3).

However, with a variant is twice as transmissible than delta and when vaccine efficacy against infection is reduced to 55%, there are large outbreaks in the university setting even at very high (90%) vaccine uptake. These outbreaks occur regardless of uptake of asymptomatic testing and peak near the end of term, with between 1300-2977 cases per day at the peak (Figure 6b), assuming no there is no importation of infection from the community. These figures are approximately 10 times greater than the peak that is seen with delta when a background rate of importation from the community is included. If vaccine escape is reduced (efficacy against infection increases to 70%) for this more transmissible variant (cf. 80% for delta), at very high levels of vaccination uptake (90%), together with natural immunity in 20% of students and no importation from the community, we still see growth in the number of cases over the course of a term (Figure 6a) rather than the decay in the number of cases that is seen in the same vaccination uptake scenario with delta (Figure 5b). However, this could be mitigated with at least 50% uptake of asymptomatic testing, which keeps the number of daily cases at a constant level.

**Figure 6.**
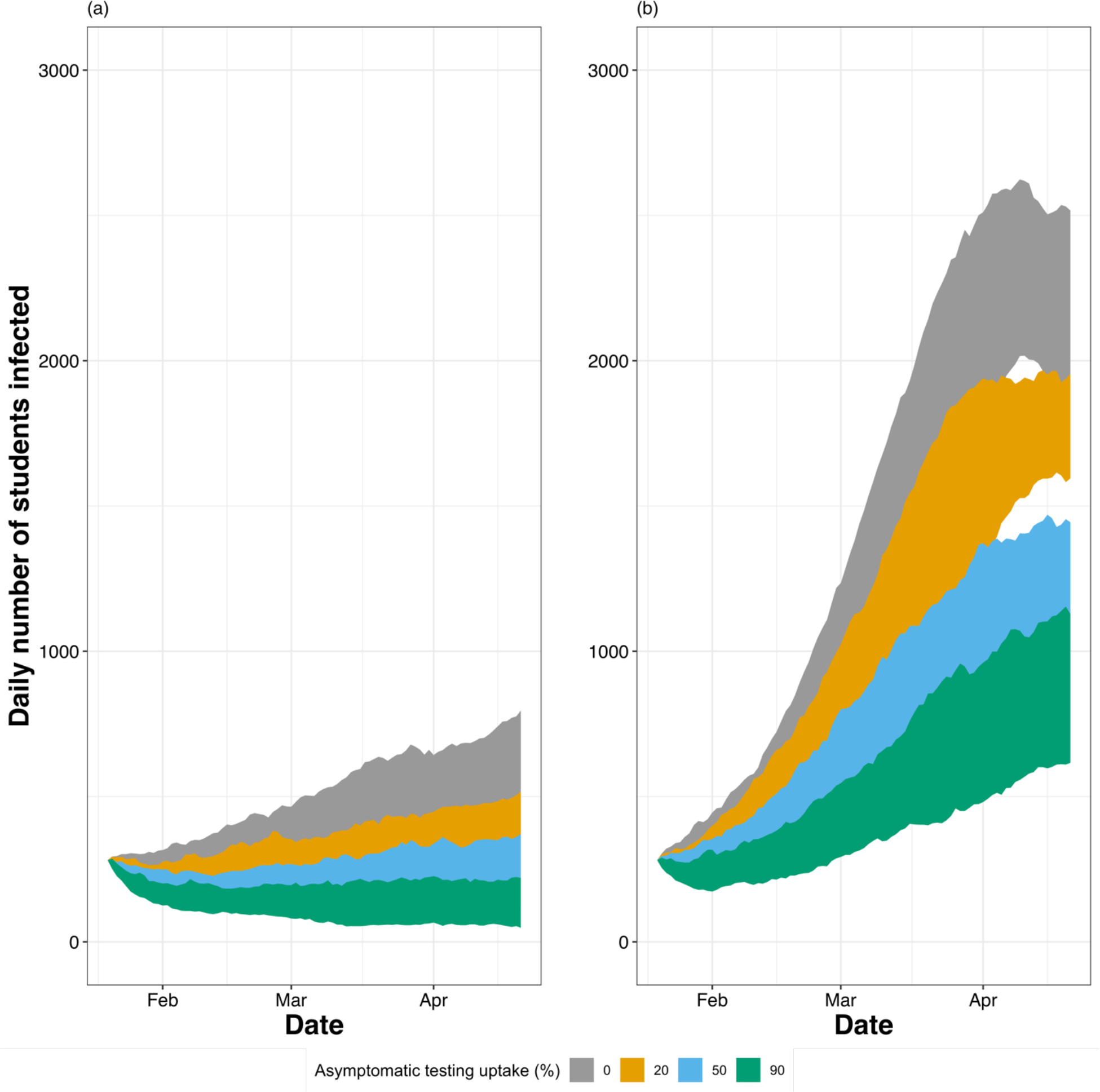
Daily numbers of SARS-CoV-2 infections in a population of students (n=28116) during the spring term of 2021 in model simulations where 90% of students are vaccinated under different levels of asymptomatic testing where a new variant is dominant that is more transmissible than delta and more able to escape vaccines. **(a)** The variant is twice as transmissible than delta and vaccine efficacy against infection by the new variant is 70% **(b)** The variant is twice as transmissible than delta and vaccine efficacy against infection by the new variant is 55%. It is assumed those who are vaccinated have a mRNA vaccine. Importation from the community is not included in these scenarios. Simulations were initialised with 1% of students infected and 20% having natural immunity. Results of the given scenarios are shown for 100 stochastic repeats.

We observed little impact of waning immunity on infection dynamics over the period studied when considering a range in the duration of protection (6,7,8 and 9 months) induced by natural infection against infection with delta (Supplementary materials).

## Discussion

We demonstrate here that SARS-CoV-2 outbreaks are very likely to occur in universities during the 2021 Autumn term where there are low to medium (30-50%) levels of vaccination uptake. With only 30% uptake of vaccination and no asymptomatic testing, these outbreaks could lead to 53-71% of students becoming infected over the course of the term depending on the number of students initially infected at the start; more than double the proportion of students estimated to have been infected in the Autumn term of 2020, when there were more social restrictions in place^21^. This may be due to the higher transmissibility of the delta variant, the strain most commonly in circulation in Autumn 2021, estimated to be 2.4 times more transmissible than the wild-type SARS-CoV-2 strain that was dominant in Autumn 2020^22,23^. At high to very high (70-90%) levels of vaccination uptake, cumulative incidence in the Autumn term of 2021 is estimated to be much lower, at 7-9%, indicating that COVID-19 vaccination uptake in a university setting is vital for controlling outbreaks. In a scenario where the dominant variant is at least twice as transmissible as delta and vaccine efficacy against infection is reduced to 70%, at least 50% uptake of asymptomatic testing is needed on top of 90% vaccination uptake to control outbreaks in a university setting. Without additional measures, large outbreaks may be seen if vaccine efficacy against infection is 55% or less.

The uptake of vaccination is expected to be variable across universities which is why we explored a range of values here. Vaccination uptake in the 18 to 24 age group (representative of most students) was estimated to be around 65-68% on the 4th of October in England for the first dose and 55-56% for the second dose^11^. However, the student COVID-19 insights survey^24^ suggests that uptake could be even higher among university students compared to others within the same age group. From 27th September to 4th October 2021, 90% of university students surveyed in England (n=960, weighted by sex, age, and region) had already had at least one SARS-CoV-2 vaccine, with most having had two doses (78%). Therefore, our scenarios with medium to high vaccination uptake (50-70%) may be the most relevant for English universities: however, the survey results are taken as an average across institutions and inevitably there will be heterogeneity in vaccine uptake across institutions. In addition, vaccination programs vary between countries, in the vaccine products used, the timings and the levels of uptake, and so our scenarios with lower vaccine uptake may be useful for informing similarly structured universities in other regions.

Our work suggests that asymptomatic testing is a useful supporting measure and can help to further reduce case numbers, having the most impact at low to medium levels of vaccination uptake. This corroborates previous findings reporting that asymptomatic testing is more impactful at higher reproduction numbers^8-10^. Reliably estimating uptake of asymptomatic testing is challenging. Although recommended by the UK Department of Education (DfE) ^5^, universities do not have the autonomy to mandate this and the uptake in students may be variable across institutions. The student COVID-19 insights survey from 27th September to 4th October 2021^24^ found that 55% of students surveyed in England had taken a COVID-19 test in the past seven days (n=960, weighted by sex, age and region), but did not distinguish between symptomatic (PCR) and asymptomatic LFT. For institutions with a low vaccination uptake, it will be particularly important to encourage asymptomatic testing uptake.

Asymptomatic testing may also be important to mitigate any increases in case numbers in the student population that may corroborate with those seen in the community. We found that under high levels of vaccination uptake (90%) that outbreaks in the university population did not occur if the university student population is “closed” (no transmission occurs between university students and other individuals). However, when importation of COVID-19 from the community into the student population does occur at a time-varying rate, with higher numbers of cases in the community expected in the winter, this can lead to an increase in student cases later in the term. The extent at which bi-directional transmission of COVID-19 occurs between the student population and the local community is unknown and is likely to vary between institutions. Previous work using age-stratified and geographical spread analyses found signals for spillover of COVID-19 from universities to the community in some local tier local authorities in England, but not in others. However, the analysis was not able to identify specific risk factors for transmission between universities and the community^8^. Therefore, our work cannot give any exact predictions or quantitative estimates for the impact of importation of COVID-19 from the community into the student population. However, our work highlights the importance for universities to be observant of the number of cases in the local community and qualitatively suggests that use of asymptomatic testing can help to mitigate any importation that may occur, even under high levels of vaccination uptake (90%).

As well as the addition of the impact of vaccination to the Brooks Pollock et al.^9^ model, we also incorporated waning immunity. However, we observed little impact of waning immunity following infection on infection dynamics over the time period studied when delta is the dominant strain. Modelling work has shown that the choice of the waning mechanism affects the dynamics of a model, both quantitatively and qualitatively, for example, through the function chosen to model the decay in immunity^25^. Here, we assumed protective immunity following natural infection to be sustained for 6 to 9 months^14,26^. Accordingly, we did not observe waning immunity to impact on infection dynamics over the 3-month period studied. Future modelling scenarios should consider including waning of vaccine efficacy and the interaction between immunity acquired through vaccination and infection.

A further extension of the model may be to include variable uptake of vaccination across school and year groups if relevant data were available. We assume in all scenarios that uptake is consistent across all faculties and year groups and that a high efficacy vaccine (such as an mRNA product) is used exclusively. However, there may be clusters of students within faculties who are not vaccinated or faculties with high representation of international students who may have been offered a different vaccine, possibly with a lower efficacy. In the model we use here, this could have implications for the mixing within and between our school/year groups. However, we did not have access to data on the vaccination status of students at this group level. Network modelling of influenza in a US high school has shown that the mean outbreak size is usually larger for networks where unvaccinated individuals have more contact with other unvaccinated individuals (positively assortative networks) compared to unvaccinated individuals contacting others at random, regardless of their vaccination status ^27^. Therefore, we may see more cases than estimated here if vaccine uptake or efficacy is unevenly distributed across faculties.

In the modelling scenarios presented here, we have not incorporated the impact of physical distancing or use of face-coverings since these were not legal governmental requirements during the period studied. However, this may have led to overestimates of outbreak sizes and case numbers in our results, since these measures are still recommended in many institutions. The student COVID-19 insights survey from 27th September to 4th October 2021^24^ found that 49% of university students surveyed in England (n = 960, weighted by sex, age and region) were trying to keep a 2-metre distance from people outside their household always or most of the time. However, this is a small sample size that may not be representative of all university settings and there may not always be opportunities for physical distancing even if students are compliant with recommendations.

Due to the primarily young demographic of the university student population, the majority (∼80%) of COVID-19 infections that occur will be expected to be asymptomatic^6,7^ and few are expected to lead to hospitalisations and deaths^28^. However, infections within a university population can pose a risk for vulnerable students and other vulnerable individuals that students may encounter, such as family members. There has been little work to quantify the frequency of long-lasting clinical sequelae resulting from COVID-19 infection (“long COVID”) in this age group^29^, however, Office for National Statistics survey data estimates^30^ suggest that there were 142,000 (95% confidence interval= 125,000-159,000) young people aged 17 to 24 in the UK with self-reported long covid on the 4th November 2021. Therefore, trying to reduce case numbers in the student population is important for protecting the vulnerable and for reducing the likelihood of students having long lasting clinical sequelae from COVID-19 infection.

In conclusion, the presence, magnitude, and timing of COVID-19 outbreaks in universities is highly dependent on the level of vaccination and asymptomatic testing uptake. Asymptomatic testing is particularly impactful at low levels of vaccination uptake. For a delta-like variant, medium to high levels of vaccination uptake and asymptomatic testing can mitigate outbreaks. However, with an omicron-like variant (twice as transmissible as delta and with vaccine efficacy against infection at 55%) large outbreaks are likely to occur in university settings even under very high levels of vaccination uptake and asymptomatic testing. If vaccine escape is less prominent, with 70% of infections being prevented by vaccination, then high uptake of asymptomatic testing (50%+) and vaccination (90%) is needed to control outbreaks of an omicron-like variant in a university setting.

## Methods

The extent and nature of outbreaks of COVID-19 in the student population during the autumn term of 2021/2022 at the University of Bristol (UOB) is investigated by incorporating vaccination and waning immunity from natural infection into an existing stochastic compartmental model of COVID-19 at UOB^9^, updating the parameters and modelling realistic scenarios.

### Model structure

The stochastic compartmental model contains seven disease states: Susceptible to SARS-CoV-2 (S), Exposed and latently infected (E), pre-symptomatic and infectious (P), symptomatic and infectious (I), asymptomatic and infectious (A), isolating/in quarantine (Q), recovered and immune (R). *N* denotes the total number of students in the population. Hospitalisations and deaths were not included since we expect low rates of hospitalisations in university students with the majority being young adults^28^. With the addition of vaccination, the rates of hospitalisation and death from the delta variant are thought to be reduced by around 85-99% and 89-97% after one or two doses of a mRNA vaccine respectively^31^. The flow and transitions between compartments are shown in Figure 1 and are given in Equations 1 and 2.

The model is structured into 161 groups, each of which represents a school within the university and a year group (for example, one group could be first year undergraduates in the School of Biological Sciences). The school/year groups were established using an anonymised extract of student data for a university from 2019/2020 which included data on primary faculty affiliation (7 faculties), primary school affiliation (28 schools), age, year of study (6 undergraduate years, taught postgraduates, and research postgraduates), home region (UK students only) and country of origin and term-time residence. The study complied with the university data-protection policy for research studies (http://www.bristol.ac.uk/media-library/sites/secretary/documents/information-governance/data-protection-policy.pdf).

The household contact rates between students in each school/year group was estimated using the term-time residence postcodes listed in the university data as described in Brooks Pollock et al.^9^. We assume that students in university accommodation would not have high rates of contact with more than 24 individuals within their residence. If the number of students at a single postcode exceeded 24, then subunits of 24 or fewer individuals were created within the postcode. Study and leisure/other contacts between groups were estimated using data from participants who listed their occupation as “STUDENT” (n=363) in the Social Contacts Survey (SCS) which was an online and paper-based survey carried out in 2010^32,33^. It was assumed that all study contacts were within the same school/year group, while leisure/other contacts were assumed to take place across the whole university as described in Brooks-Pollock et al.^9^

The flow between compartments is given by

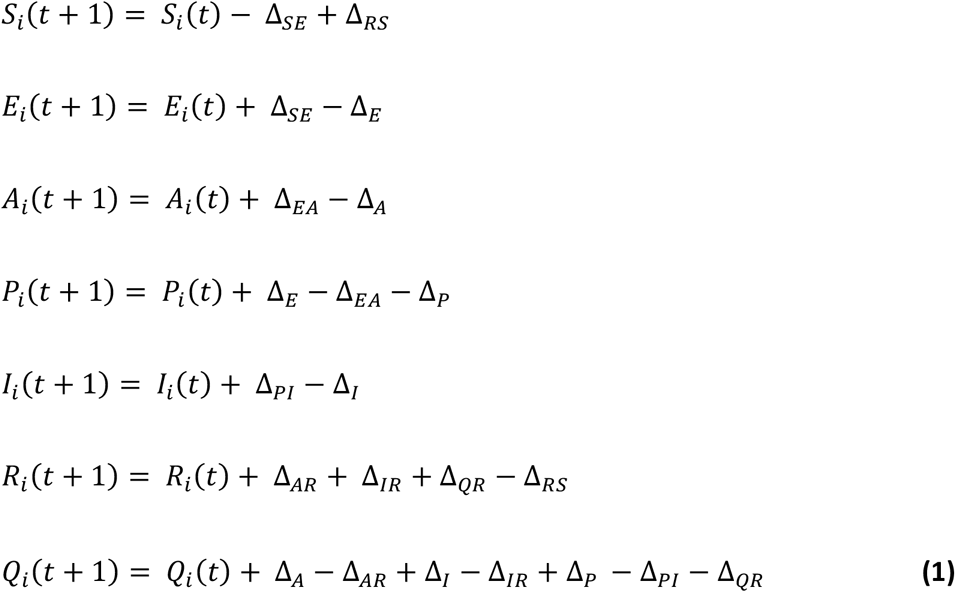

and is presented in Figure 1. The transitions between compartments are given by

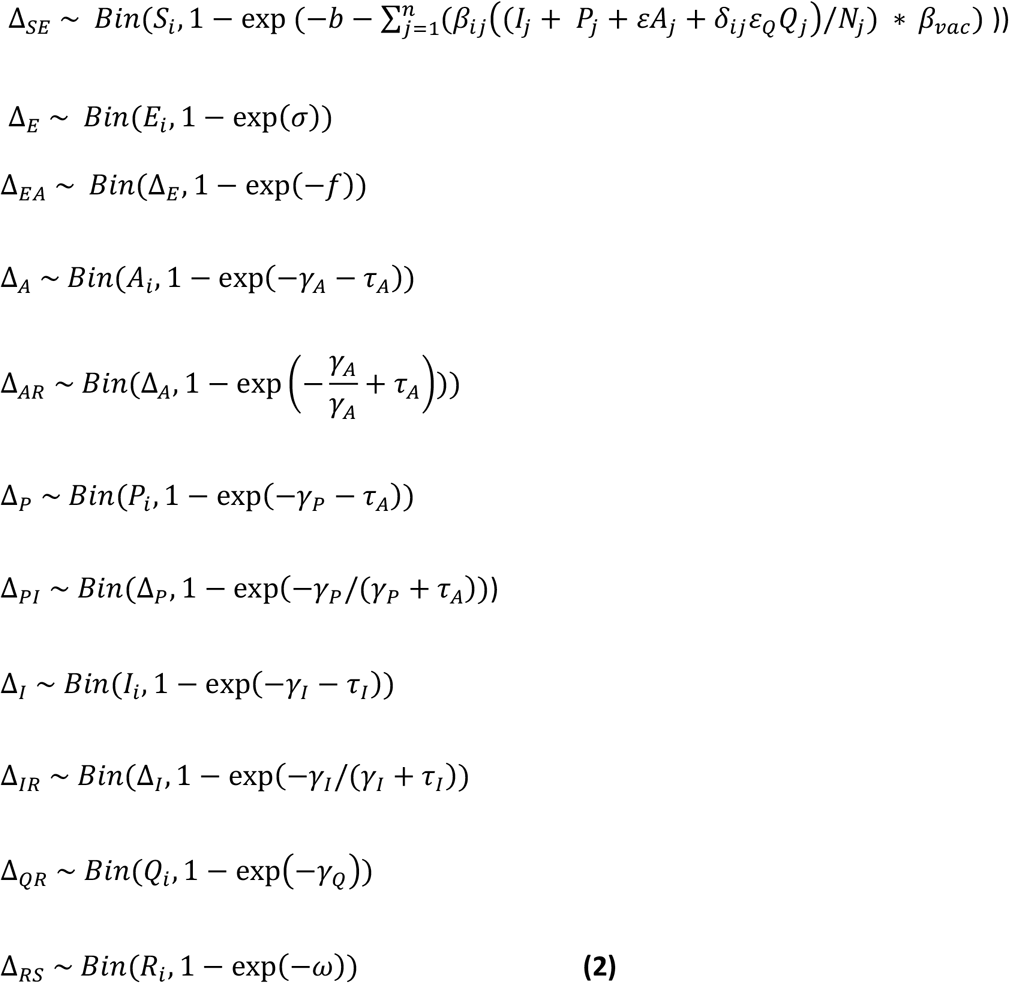

All state and transition variables are time dependent, however (*t*) is not shown in equation 2 to improve its readability. Equations 1 and 2 have been adapted from equations in Brooks-Pollock et al.^9^ to incorporate the impact of vaccination and of waning immunity from natural infection.

### Incorporating vaccination into the model

Vaccination was not included in the original model by Brooks-Pollock et al.^9^. Here, we incorporate vaccination by assuming that vaccine uptake is equal across all years and schools. This simplifying assumption means that the transmission rate in the baseline model, *β*, is scaled by a factor (*β*_*vac*_), as shown in equation 3 *β*_*vac*_ is dependent on vaccine uptake in the student body, *p*_*v*_, the relative susceptibility of vaccinated individuals, *v*_*s*_ and the relative infectiousness of vaccinated individuals, *v*_*t*_.

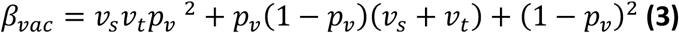

We also have scaled *f*, the proportion of cases with no symptoms, “asymptomatic cases”, according to the efficacy of vaccination reducing symptoms, *δ* and the levels of vaccination uptake in the student body *p:*

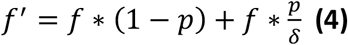

We look at different levels of vaccination uptake in the student population, assuming full protection from two doses of a mRNA vaccine. Although this may not be true of all students, due to the age demographics of the student population, these are the most likely products that will have been used. We use the estimated vaccine efficacy on reducing transmission (by 45%), susceptibility (by 80%) and symptoms (by 84%) against the delta variant, as used in the roadmap modelling by Warwick^31^ when looking at the delta variant. We reduce the vaccine efficacy on reducing transmission to 70% and then 55% when looking at the omicron variant.

### Incorporating waning immunity into the model

Waning immunity from natural infection is incorporated by the addition of a transition from the R to S compartment (Δ_*RS*_), with the assumption that loss of protection against re-infection occurs 8 months following recovery from SARS-CoV-2 infection 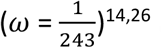. A sensitivity analysis on this parameter was performed on delta scenarios and the results are included in the Supplementary material. In the absence of data, we assumed that individuals who have natural immunity from natural infection would be as protected from the omicron-like variant as those challenged with delta.

### Testing

We assume that it takes 48 hours following symptom onset for symptomatic cases to be moved into isolation and that all are tested using a polymerase chain reaction (PCR) test. Mass testing of asymptomatic cases using LFT is another measure that has been used by universities to help mitigate the impact of COVID-19. The UK Department for Education have recommended that students should have 2 LFT per week, 3 to 4 days apart^5^. In the model scenarios which include mass testing using LFT, the test rate for asymptomatic cases is determined by the average time to test for asymptomatic cases (4 days, 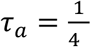) and scaled by the sensitivity of LFTs, which is estimated to be 58% for asymptomatic cases^34^ and the uptake of asymptomatic testing in the student body, *p*_*a*_.

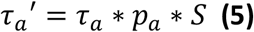

If asymptomatic cases are tested positive, they are moved to self-isolation and remain there for an average of 10 days. As the current levels of this asymptomatic testing regime are uncertain, we look at different scenarios of asymptomatic testing uptake in the student body to see what the impact would be if these levels of uptake could be encouraged.

### Isolation

Quarantining is included with symptomatic individuals being detected by PCR tests and asymptomatic cases being detected with LFT. Students who isolate in the model (are in the ‘Q’ disease state) and contribute to the force-of-infection within their school/group at a reduced rate (*εQ*=0.5), since it is often not possible for students to have no contacts when isolating. A longitudinal social contacts survey at the University of Bristol from September to November 2020 found that 99% of students who had a positive COVID-19 test 2 weeks prior to their competition of the survey had been isolating within the past week. However, many of them still had high numbers of contacts while isolating (mean = 4.3, standard deviation = 10.6) ^35^.

### Initial conditions

To estimate the number of students that are infected at the start of the simulations, we use estimates from the ONS infection survey that estimated between 26th September and 2nd October 2021, 1.44% (1.35%-1.53%) of people in England had COVID-19^36^. This rate was lower in people aged 17-to-24 years at 0.8%. Therefore, we considered three scenarios for initial incidence: low (0.5%), medium (1%) and high (2%). We assume that 50% of all infections are in the exposed and latently infected compartment (E) and, of the remaining 50% of infections, 80% (40% of total infections) are asymptomatic (based on estimations by Poletti et al. and Sah et al. ^6,7^ for this age group) and 20% (10% of total infections) are split evenly between the pre-symptomatic and infectious (P) and symptomatic and infectious (I) compartments. We assume that no students are isolating at the start of the simulations.

Due to the uncertainty around the number students who are protected against COVID-19 due to a previous natural infection, we used three different proportions of students (5%, 10% and 20%) who were initially in the recovered and immune disease state. These values were based upon estimates provided by the University of Bristol where 10% of students were infected in the 9 months prior to the start of the autumn term of 2021/2022 year, but not all summer infections were captured (in preparation).

The relative infectiousness of asymptomatic cases compared to symptomatic cases (*ε*) used here is 0.5, the baseline value from Brooks Pollock et al.^9^.

#### Importation of COVID-19 into the student population

The student body is not a completely closed population. A longitudinal social contacts survey at the University of Bristol from September to November 2020 found that 40% contacts reported by students were with individuals external to the university^35^. Students also come into contact with university staff. We use the estimate of an average of 50,000 daily infections in England in October to December 2021 from the Warwick roadmap modelling^31^and a denominator of 55.98 million (the population in England), giving an estimated maximum background infection rate (*b*) of 1e-3. This rate is time-varying to capture seasonal patterns where we expect to see higher rates of transmission into the student body from the community in the winter months. This is applied by multiplying *b* by the following term:

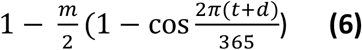

Where *t* is the timestep, *d* is the numbered day of the year for the first day of the simulation and *m* is the magnitude of the seasonal difference in transmission (where *m*=0 indicates no seasonality and *m*=1 gives maximum seasonality with no transmission at the peak of the summer. We use *m*= 1 here).

#### Impact of the delta and omicron variants

The main COVID-19 variant in circulation from the 12th September to 1st October 2021 was delta (B.1.617.2)^37^. This has been found to be approximately 60% more transmissible than the alpha variant (B.1.1.7), which was identified in late 2020^22^. The original model by Brooks-Pollock et al.^9^ estimated transmission based on the wild-type SARS-CoV-2 strain, this is estimated to be 50% less transmissible than alpha^23^. Therefore, for the majority of scenarios, we have increased R to reflect the increase in transmissibility from the wild type to B.1.1.7, and then to B.1.617.2 by multiplying the transmission matrix by 2.4. When investigating an omicron-like variant, twice as transmissible as delta, we multiply this again by 2.

## Data Availability

Contact the corresponding author regarding the data used for modelling.

## Acknowledgements

We thank the University of Bristol for providing the data and the University of Bristol Scientific Advisory Group for support. Thank you to the Bristol UNCOVER group for facilitating collaborations. HC, EBP and RK would like to acknowledge support from the National Institute for Health Research (NIHR) Health Protection Research Unit (HPRU) in Behavioural Science and Evaluation at the University of Bristol. HC is additionally funded through an NIHR Career Development Fellowship [CDF-2018-11-ST2-015]. The views expressed are those of the author(s) and not necessarily those of the NIHR, the Department of Health and Social Care, or UKHSA. CR is a member of the MRC Integrative Epidemiology Unit and receives support from the MRC (MC_UU_00011/5) and the University of Bristol. ATh is supported by the Wellcome Trust (217509/Z/19/Z) and UKRI through the JUNIPER consortium MR/V038613/1 and CoMMinS study MR/V028545/1. EBP, EN, AB and LD are supported by UKRI through the JUNIPER consortium (Grant Number MR/V038613/1). LD and EBP are further supported by MRC (Grant Number MC/PC/19067). LD acknowledges funding from EPSRC (EP/V051555/1 and The Alan Turing Institute, Grant EP/N510129/1). DE acknowledges funding from EPSRC (EP/W000032/1). RK is funded by the Wellcome GW4 Clinical Academic Training programme (203918).

## Author contributions

EN, EBP and HC conceived the study. EN, EBP, ATh, HC, CR and AB designed the study. EN and EBP modified and ran the model. EN produced the figures. EN and DS wrote the first draft of the paper. EN, ATh, DS, AB, GH, AT, LD, RK, JW, DE, CR, HC and EBP interpreted the results and revised the paper.

