## Supplementa for "Impacts of vaccination and asymptomatic testing on SARS-CoV-2 transmission dynamics in a university setting" (from another version of this preprint: version 1, 2021-11-24)

### Sensitivity analysis of waning immunity

All results given in the paper are based on the assumption that waning immunity from natural infection starts at 8 months post infection. The following plots show results from all the parameter combinations used in the paper repeated for four different estimates of time until waning immunity from natural infection (6,7,8,9 months).

*Without importation of infection from the community*

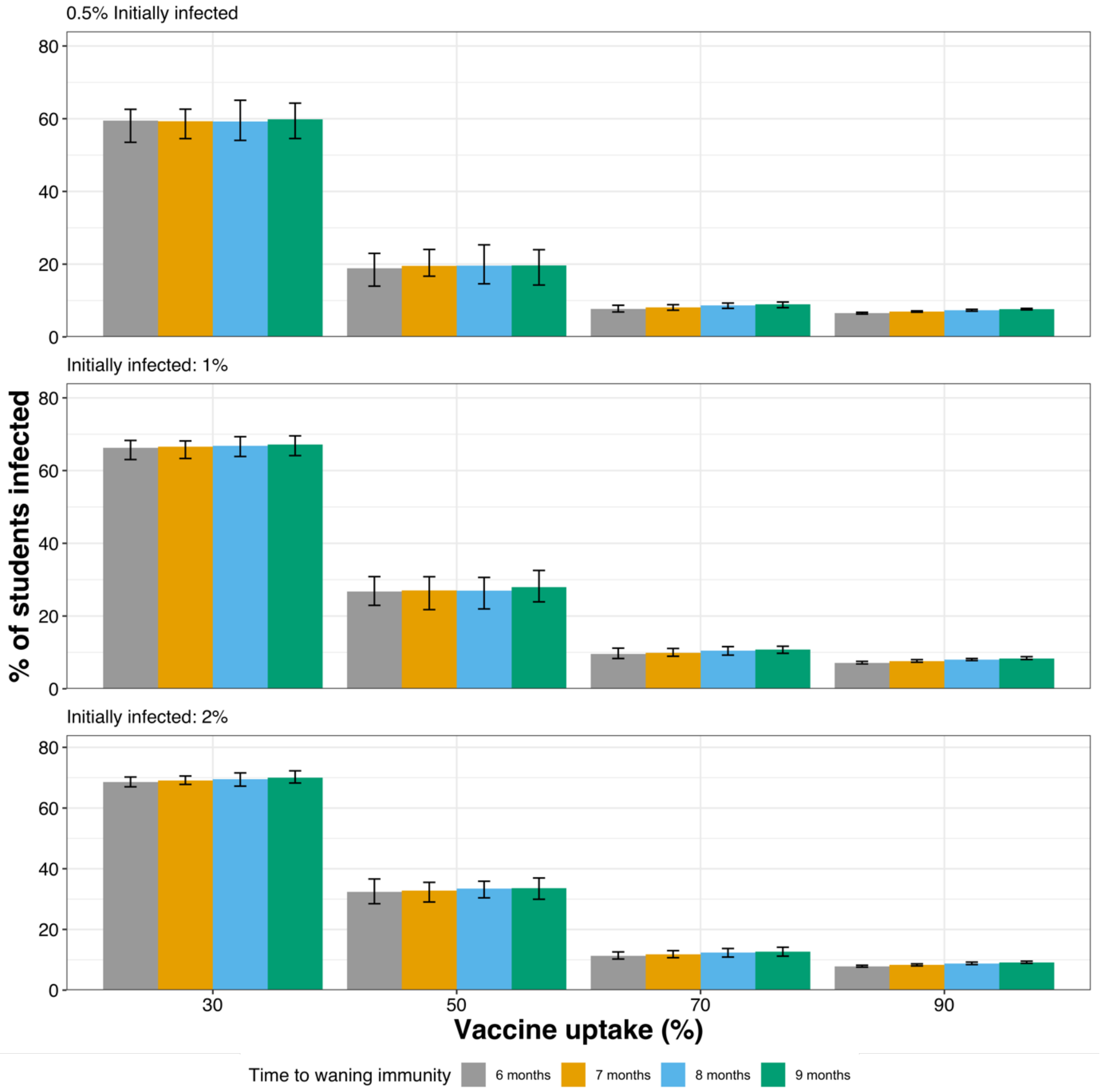


**Figure S1. The cumulative percentage of university students infected with SARS-CoV-2 in the Autumn term of 2021 as predicted under different scenarios of waning immunity, vaccine uptake, initially infected students and with no asymptomatic testing.** Each scenario is repeated 50 times, the mean and range are given. The results shown do not include those who were infected but whose immunity has waned by the end of term. It is assumed there is no importation of infection from community.

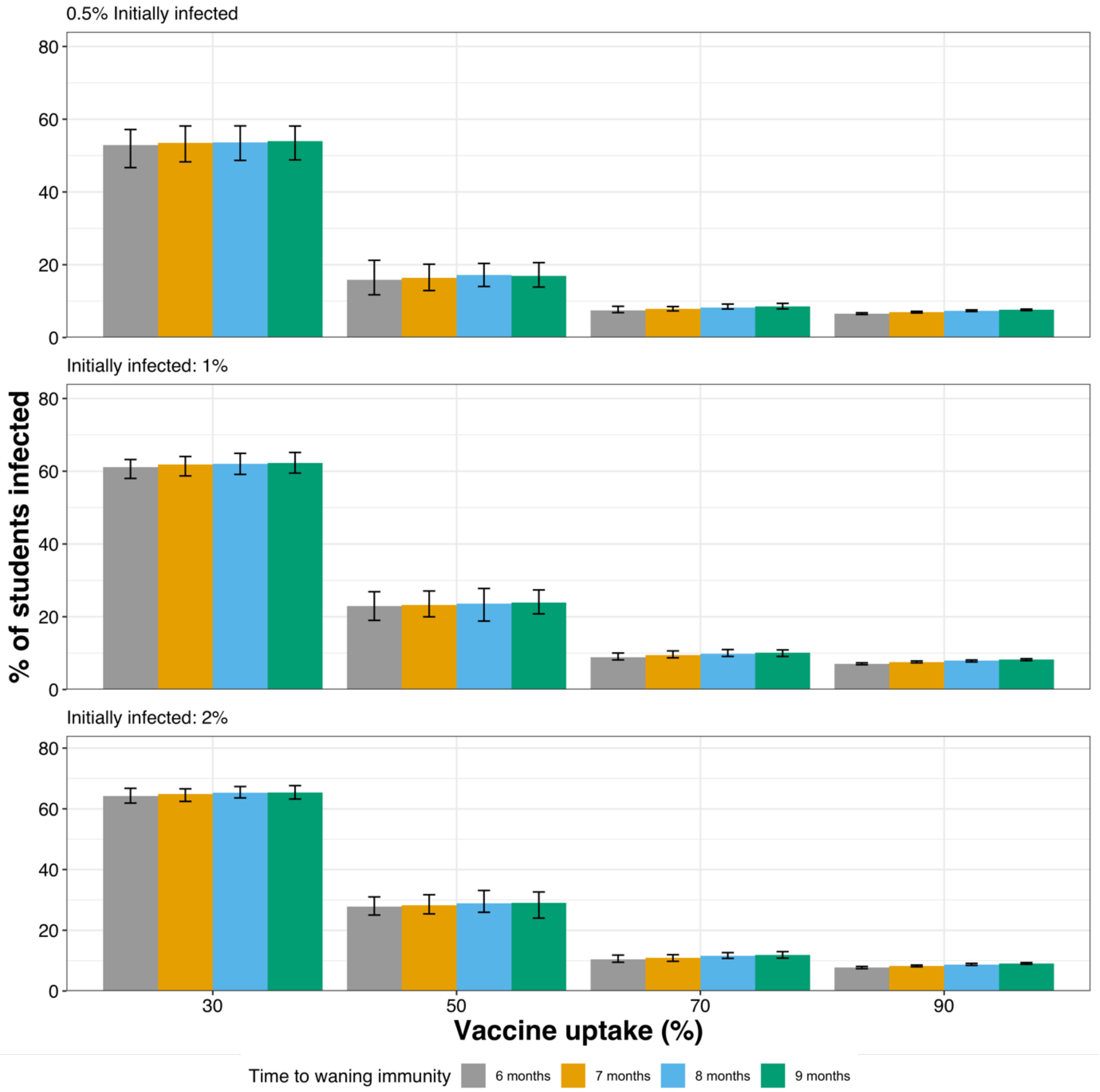


**Figure S2. The cumulative percentage of university students infected with SARS-CoV-2 in the Autumn term of 2021 as predicted under different scenarios of waning immunity, vaccine uptake, initially infected students and with 20% students taking part in asymptomatic testing.** Each scenario is repeated 50 times, the mean and range are given. The results shown do not include those who were infected but whose immunity has waned by the end of term. It is assumed there is no importation of infection from community.

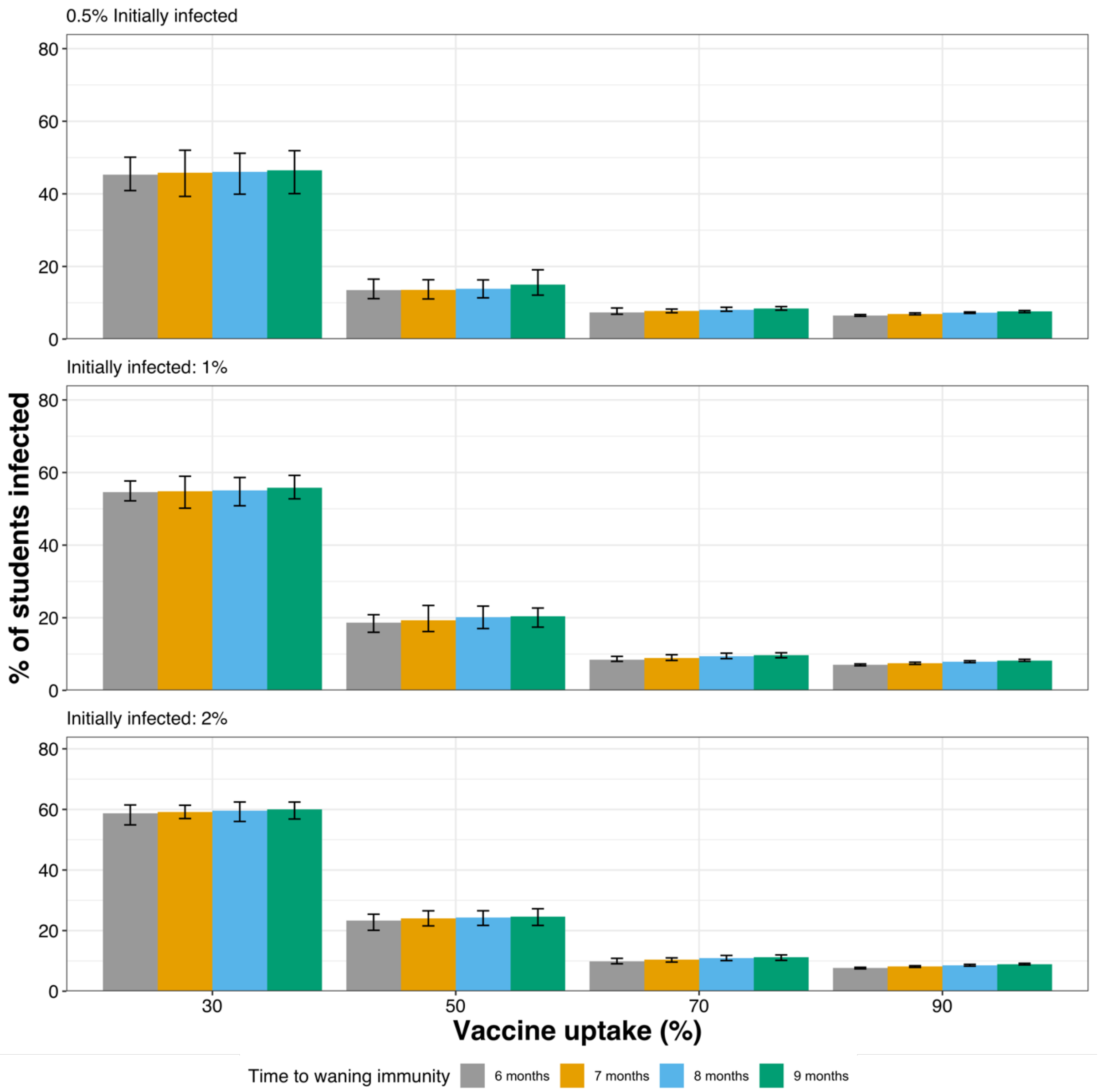


**Figure S3. The cumulative percentage of university students infected with SARS-CoV-2 in the Autumn term of 2021 as predicted under different scenarios of waning immunity, vaccine uptake, initially infected students and with 50% students taking part in asymptomatic testing.** Each scenario is repeated 50 times, the mean and range are given. The results shown do not include those who were infected but whose immunity has waned by the end of term. It is assumed there is no importation of infection from community.

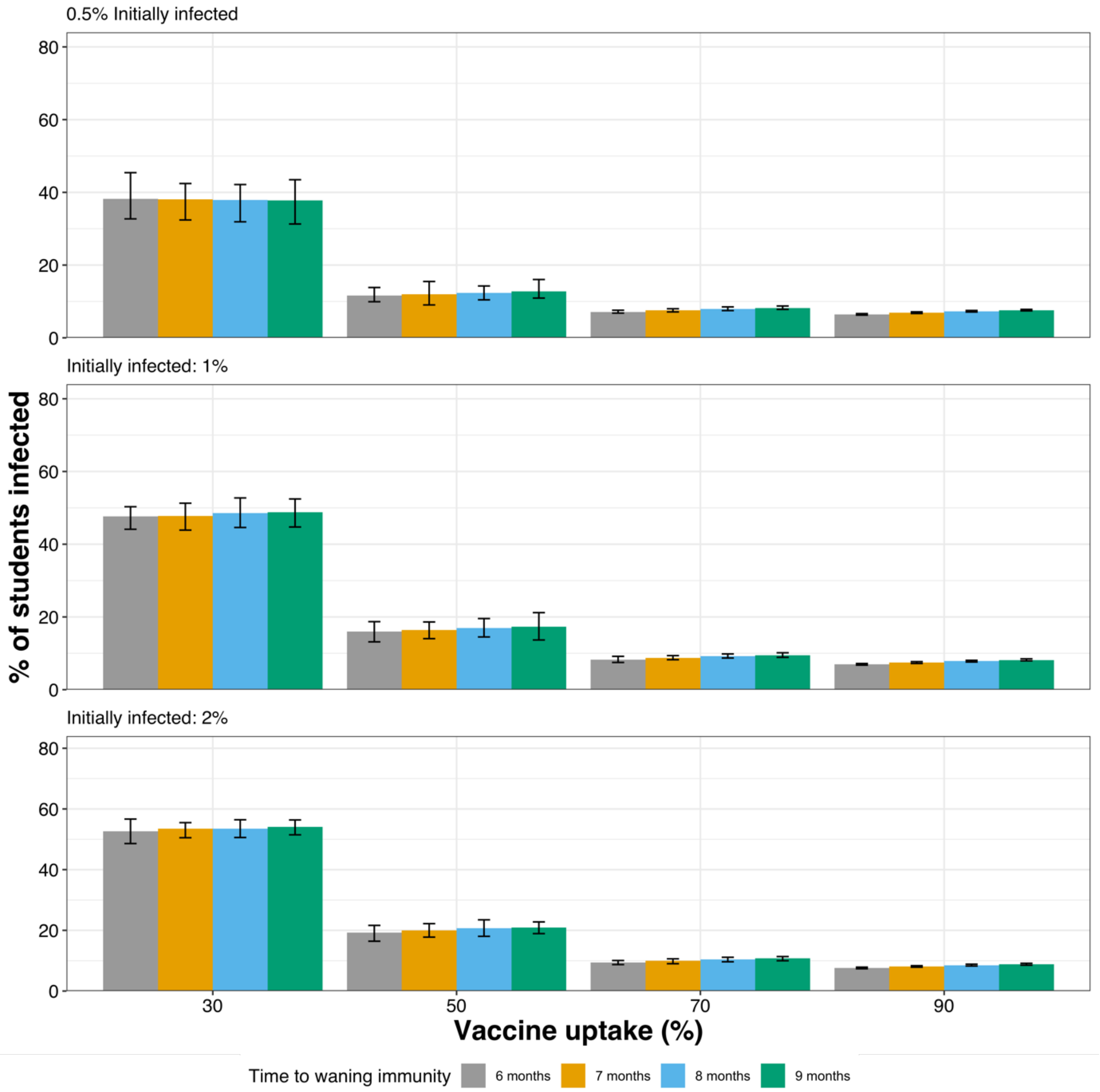


**Figure S4. The cumulative percentage of university students infected with SARS-CoV-2 in the Autumn term of 2021 as predicted under different scenarios of waning immunity, vaccine uptake, initially infected students and with 90% students taking part in asymptomatic testing.** Each scenario is repeated 50 times, the mean and range are given. The results shown do not include those who were infected but whose immunity has waned by the end of term. It is assumed there is no importation of infection from community.

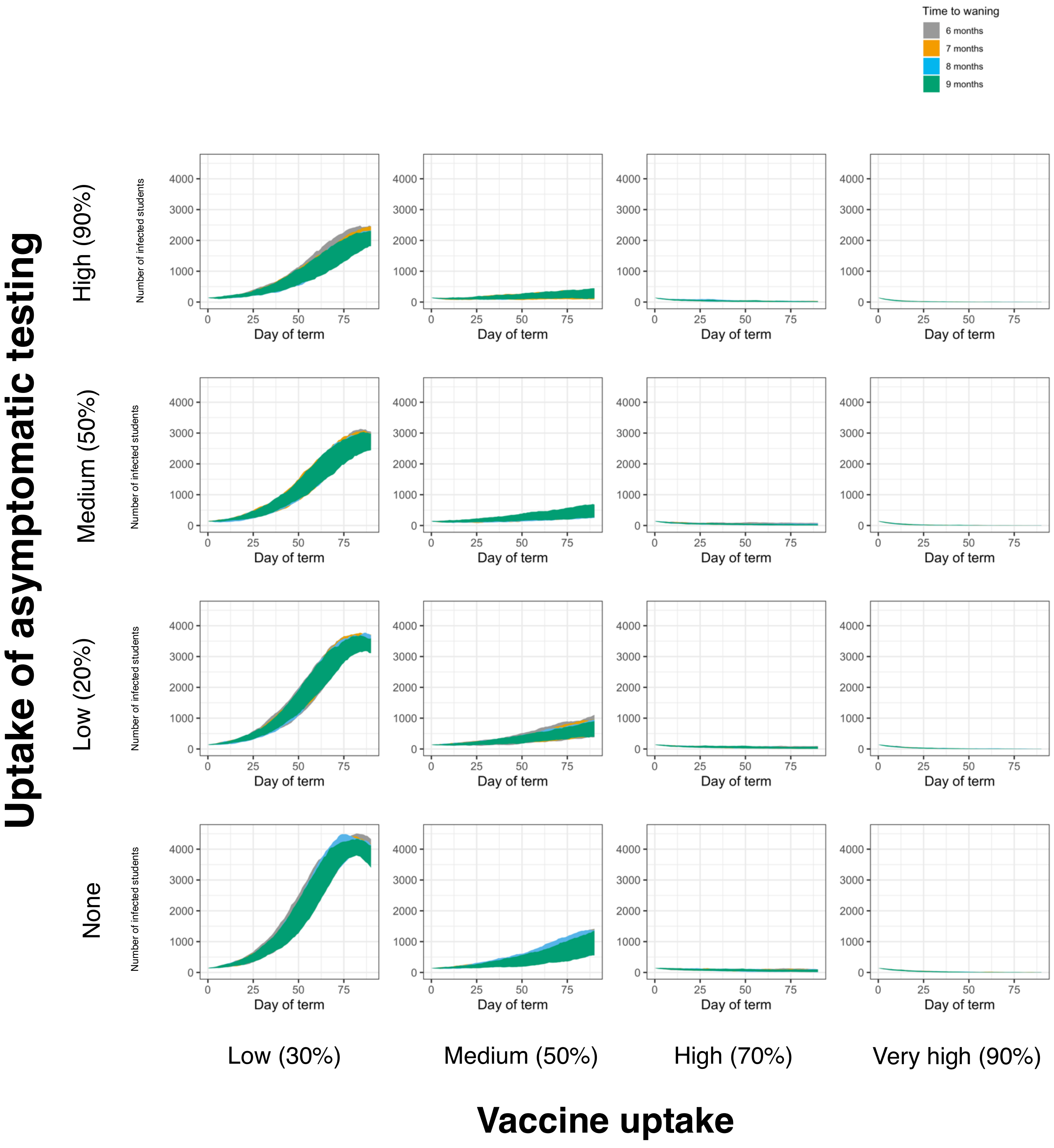


**Figure S5. Daily numbers of SARS-CoV-2 infections in a population of students (n=28116) during the autumn term of 2021 as predicted under different scenarios of waning immunity, vaccine uptake, asymptomatic testing and with 0.5% of students initially infected.** Each scenario is repeated 50 times and the range is given. It is assumed there is no importation of infection from community.

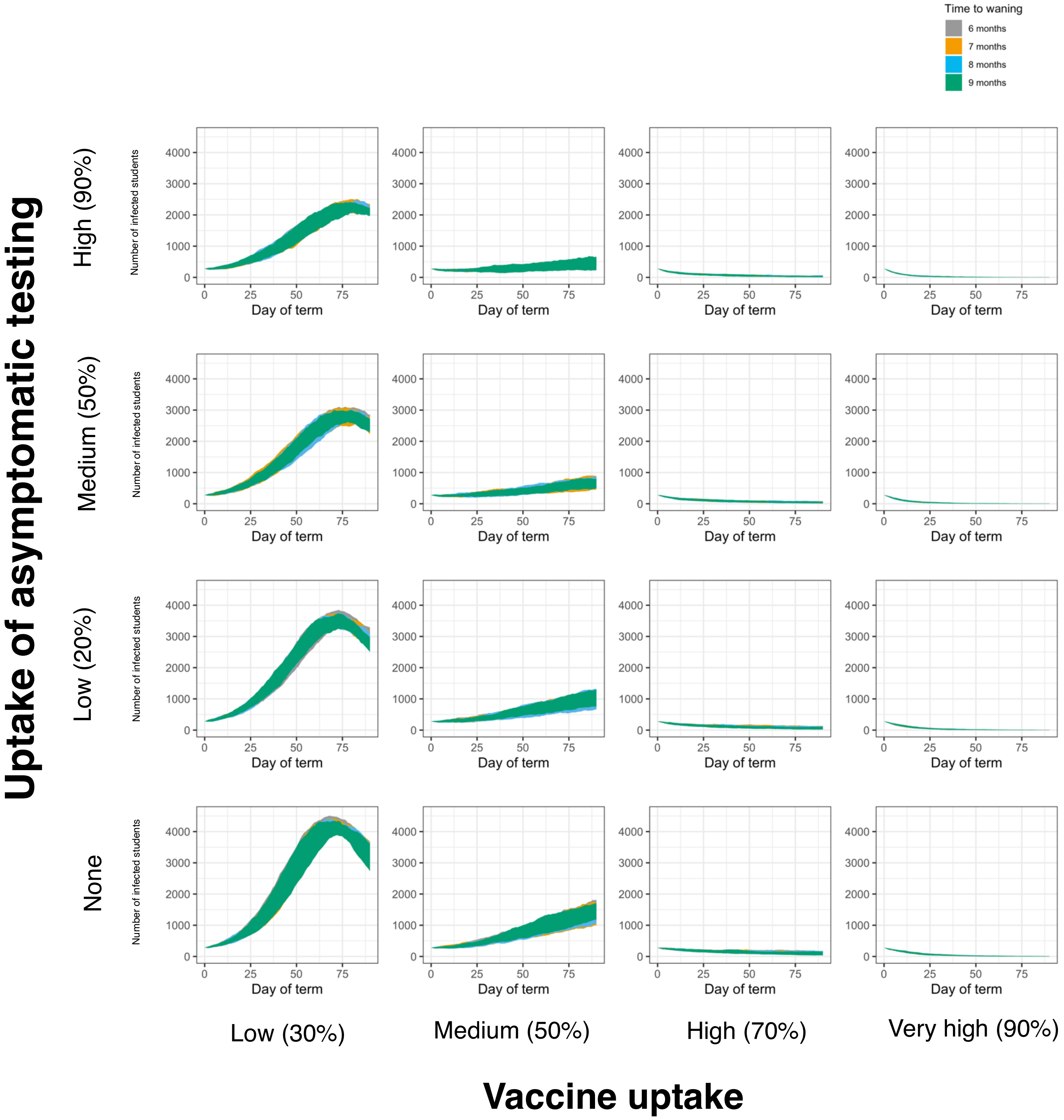


**Figure S6. Daily numbers of SARS-CoV-2 infections in a population of students (n=28116) during the autumn term of 2021 as predicted under different scenarios of waning immunity, vaccine uptake, asymptomatic testing and with 1% of students initially infected.** Each scenario is repeated 50 times and the range is given. It is assumed there is no importation of infection from community.

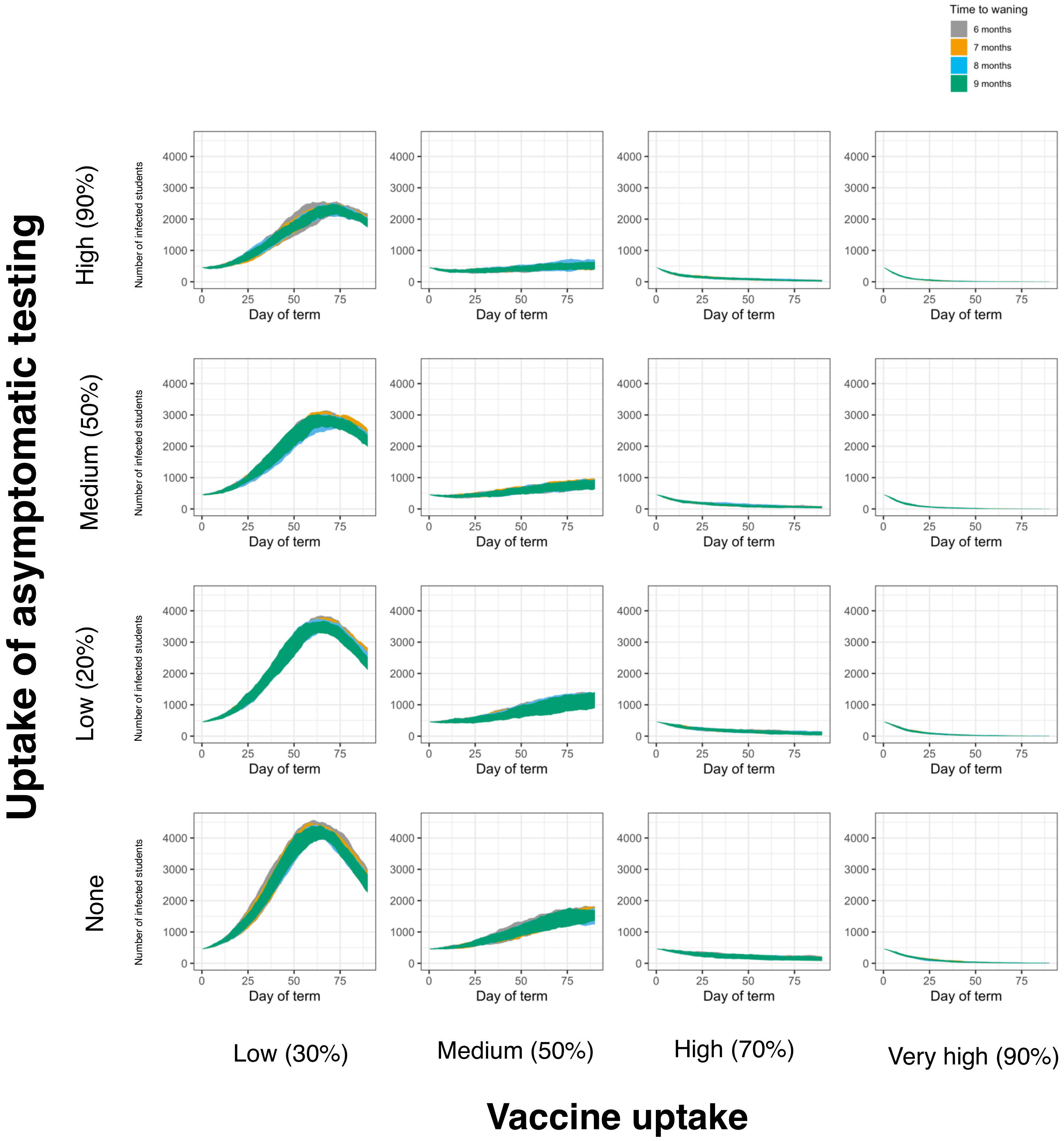


**Figure S7. Daily numbers of SARS-CoV-2 infections in a population of students (n=28116) during the autumn term of 2021 as predicted under different scenarios of waning immunity, vaccine uptake, asymptomatic testing and with 2% of students initially infected.** Each scenario is repeated 50 times and the range is given. It is assumed there is no importation of infection from community.

*With a time-varying rate of importation from the community*

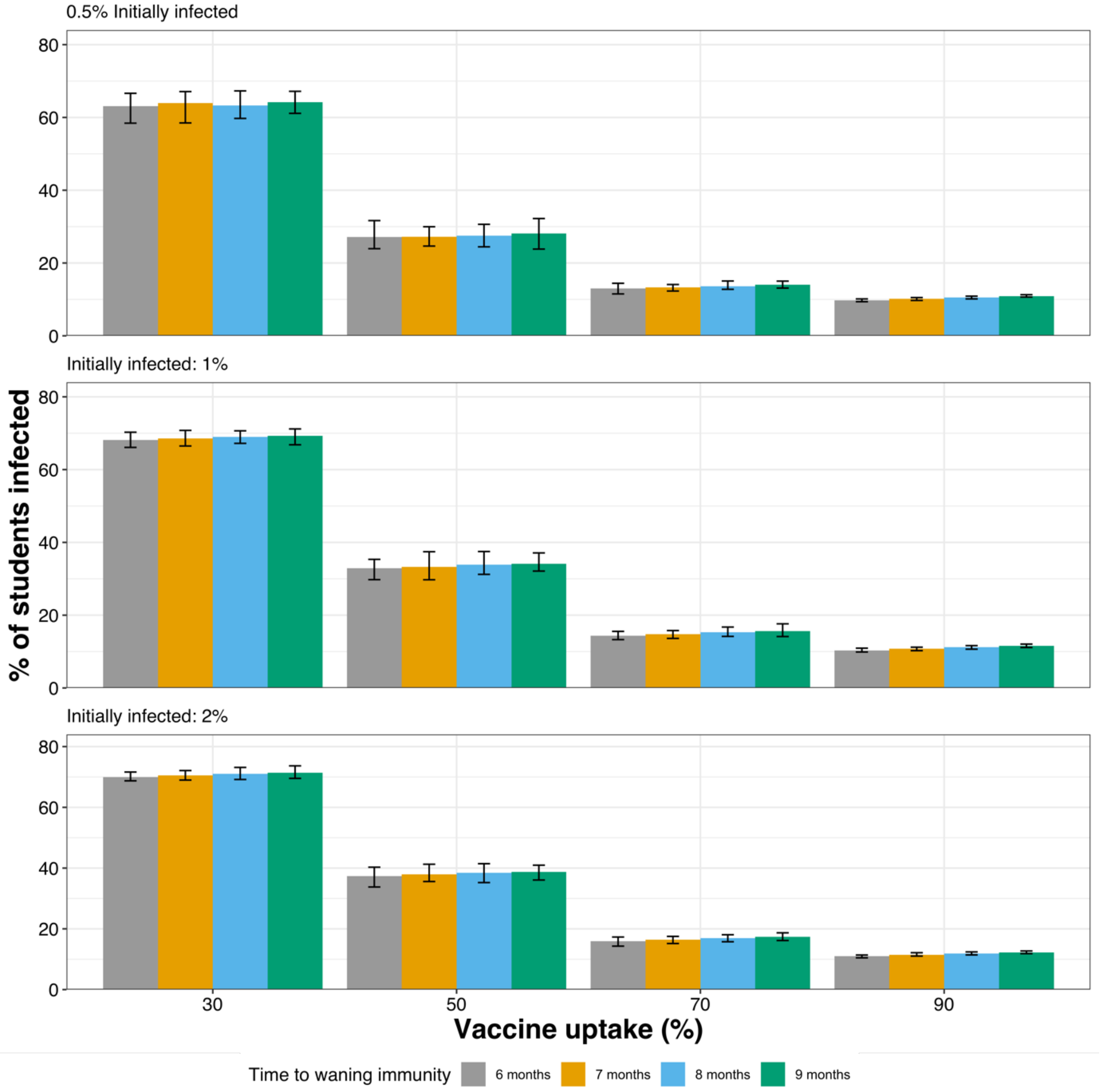


**Figure S8. The cumulative percentage of university students infected with SARS-CoV-2 in the Autumn term of 2021 as predicted under different scenarios of waning immunity, vaccine uptake, initially infected students and with no asymptomatic testing.** Each scenario is repeated 50 times, the mean and range are given. The results shown do not include those who were infected but whose immunity has waned by the end of term. Importation from the community was assumed to occur at a time-varying seasonal rate with maximum value of 1e-3.

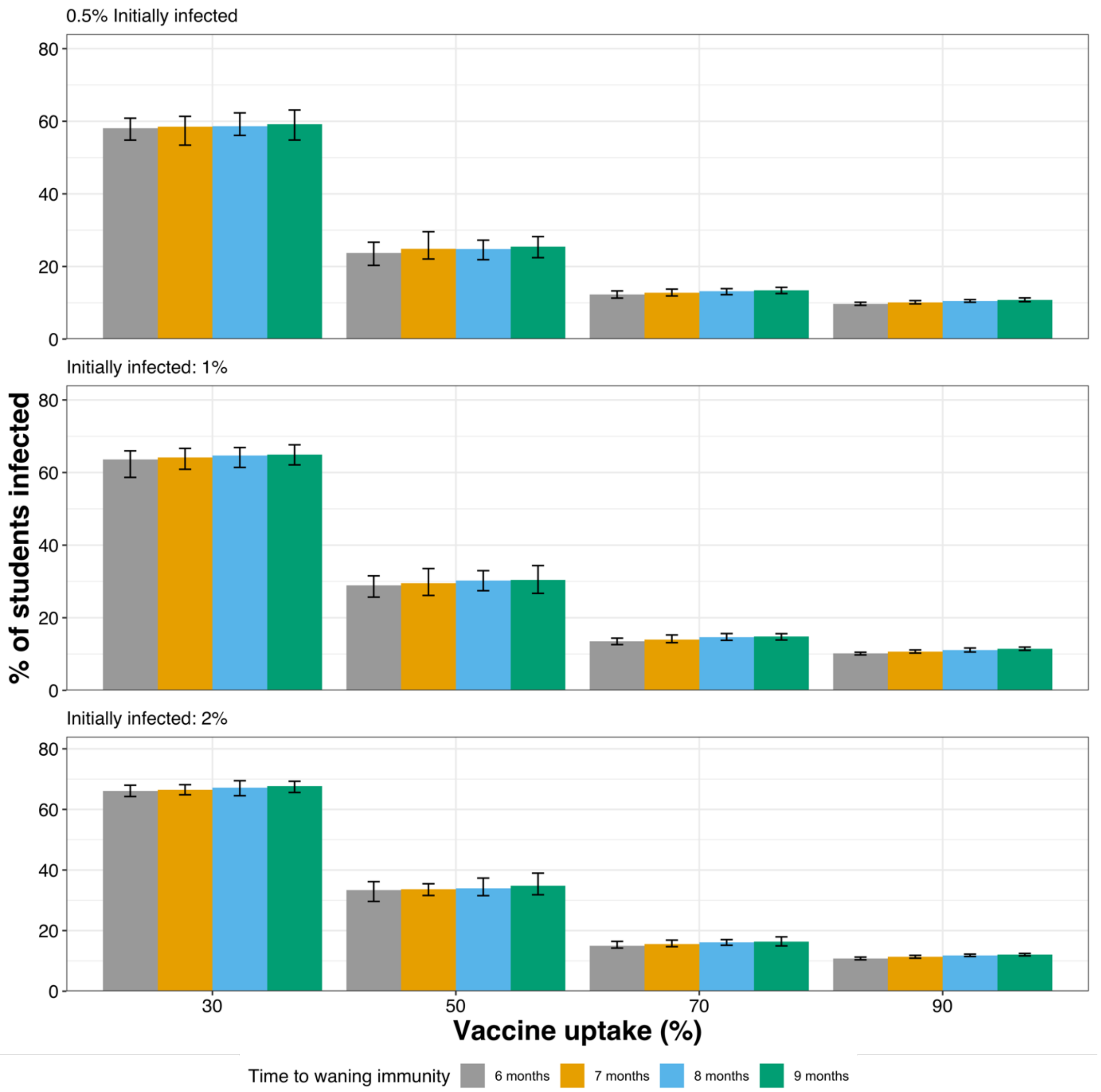


**Figure S9. The cumulative percentage of university students infected with SARS-CoV-2 in the Autumn term of 2021 as predicted under different scenarios of waning immunity, vaccine uptake, initially infected students and with 20% students taking part in asymptomatic testing.** Each scenario is repeated 50 times, the mean and range are given. The results shown do not include those who were infected but whose immunity has waned by the end of term. Importation from the community was assumed to occur at a time-varying seasonal rate with maximum value of 1e-3.

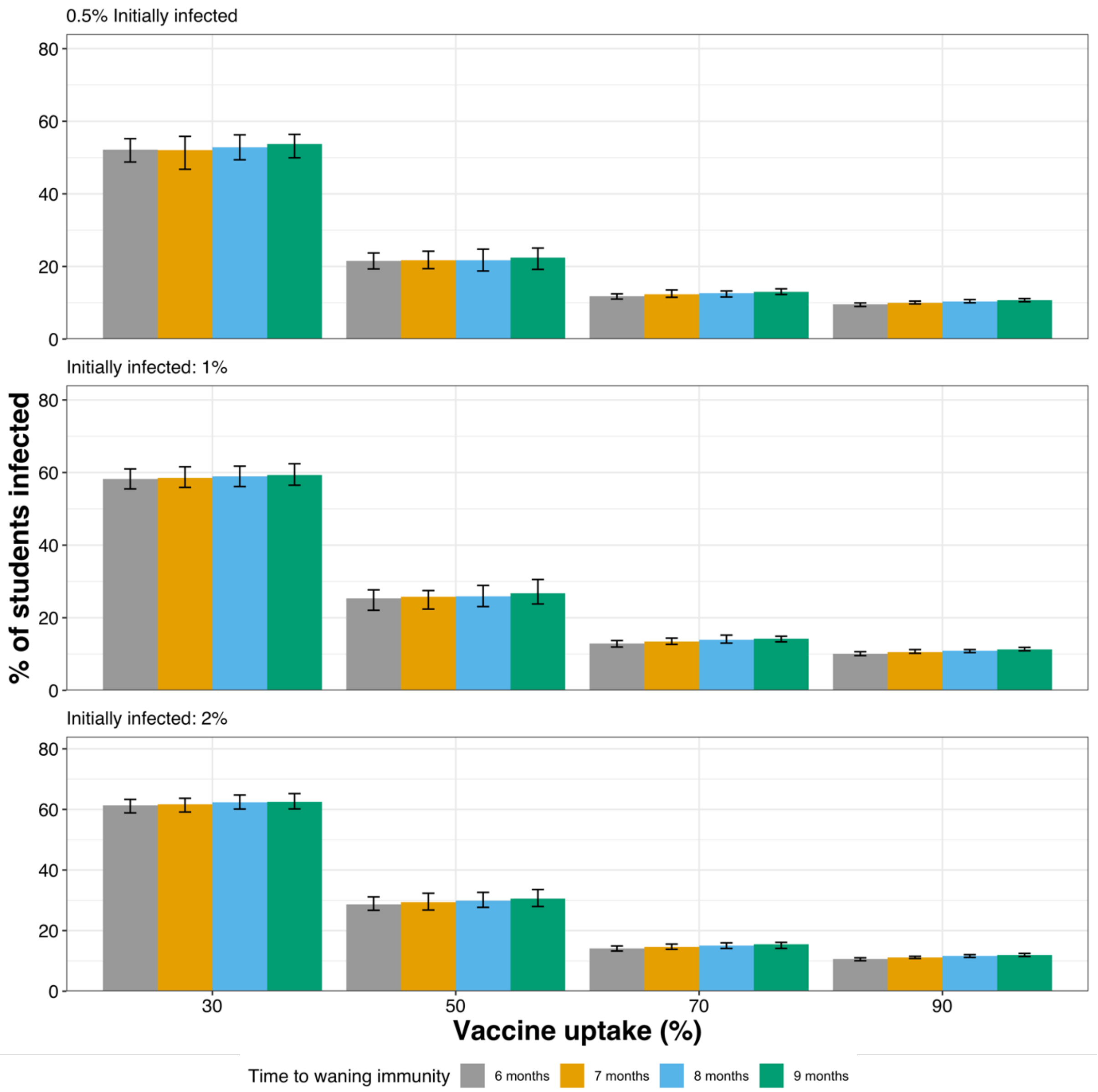


**Figure S10. The cumulative percentage of university students infected with SARS-CoV-2 in the Autumn term of 2021 as predicted under different scenarios of waning immunity, vaccine uptake, initially infected students and with 50% students taking part in asymptomatic testing.** Each scenario is repeated 50 times, the mean and range are given. The results shown do not include those who were infected but whose immunity has waned by the end of term. Importation from the community was assumed to occur at a time-varying seasonal rate with maximum value of 1e-3.

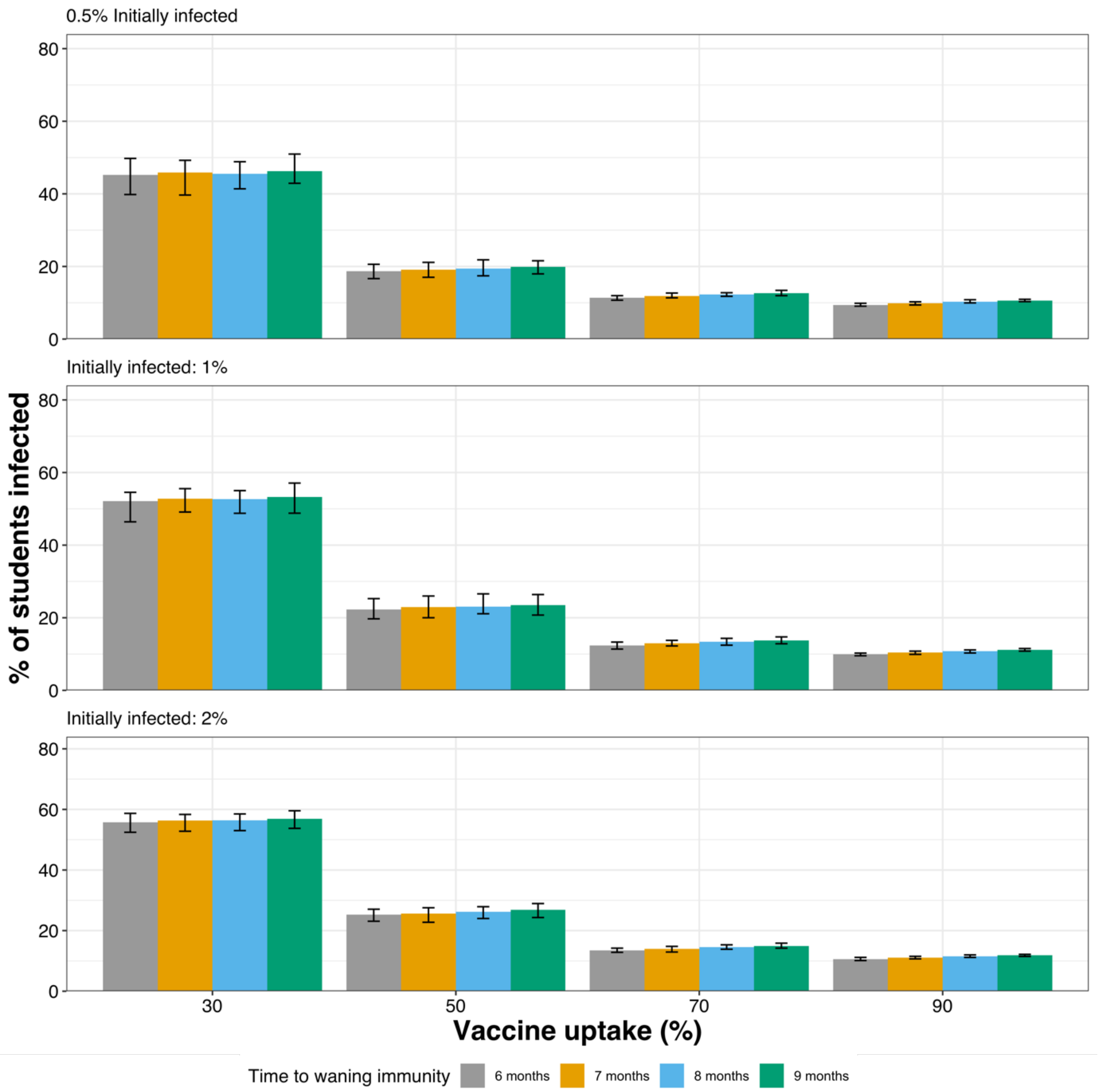


**Figure S11. The cumulative percentage of university students infected with SARS-CoV-2 in the Autumn term of 2021 as predicted under different scenarios of waning immunity, vaccine uptake, initially infected students and with 90% students taking part in asymptomatic testing.** Each scenario is repeated 50 times, the mean and range are given. The results shown do not include those who were infected but whose immunity has waned by the end of term. Importation from the community was assumed to occur at a time-varying seasonal rate with maximum value of 1e-3.

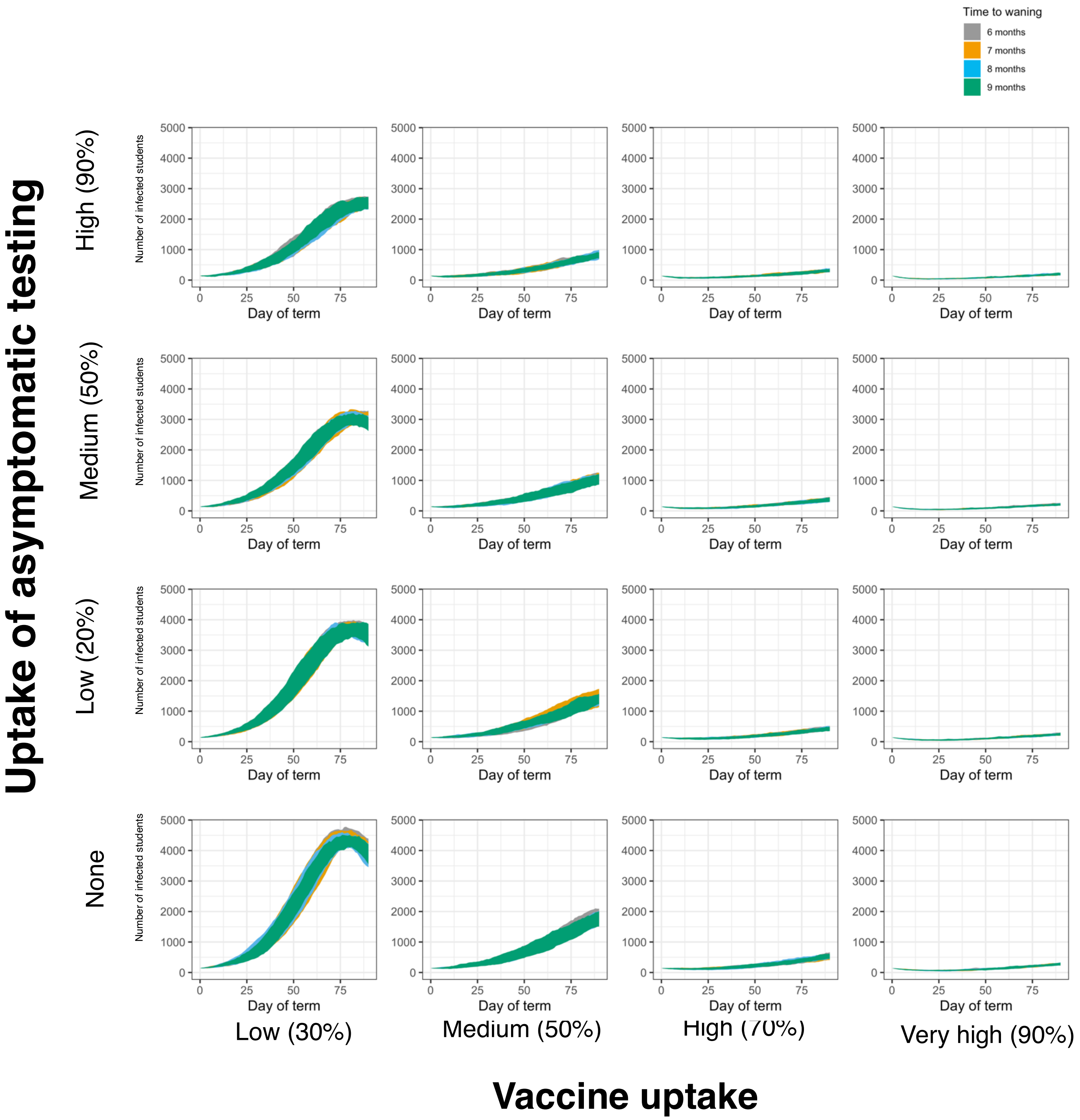


**Figure S12. Daily numbers of SARS-CoV-2 infections in a population of students (n=28116) during the autumn term of 2021 as predicted under different scenarios of waning immunity, vaccine uptake, asymptomatic testing and with 0.5% of students initially infected.** Each scenario is repeated 50 times and the range is given. Importation from the community was assumed to occur at a time-varying seasonal rate with maximum value of 1e-3.

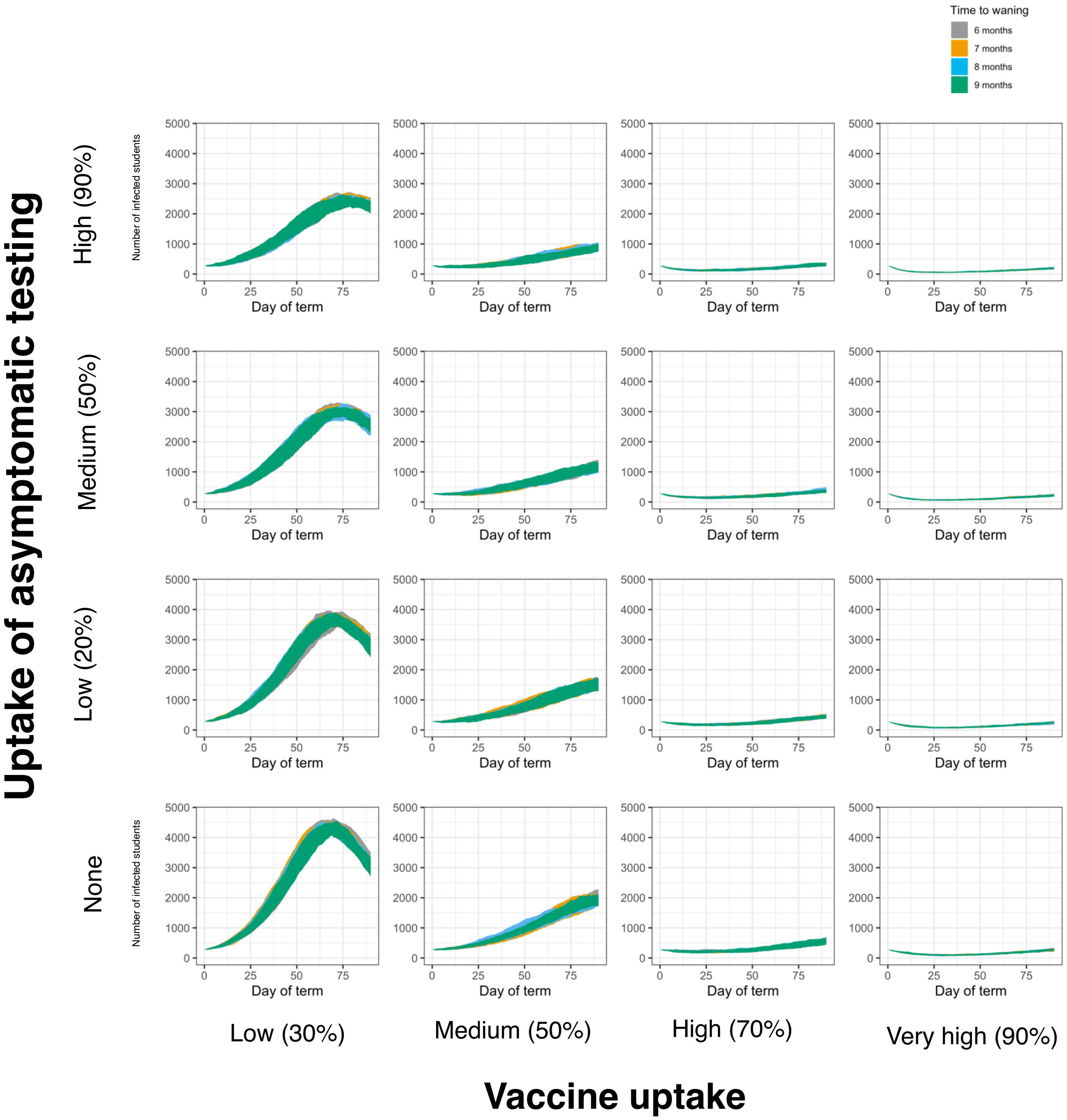


**Figure S13. Daily numbers of SARS-CoV-2 infections in a population of students (n=28116) during the autumn term of 2021 as predicted under different scenarios of waning immunity, vaccine uptake, asymptomatic testing and with 1% of students initially infected.** Each scenario is repeated 50 times and the range is given. Importation from the community was assumed to occur at a time-varying seasonal rate with maximum value of 1e-3.

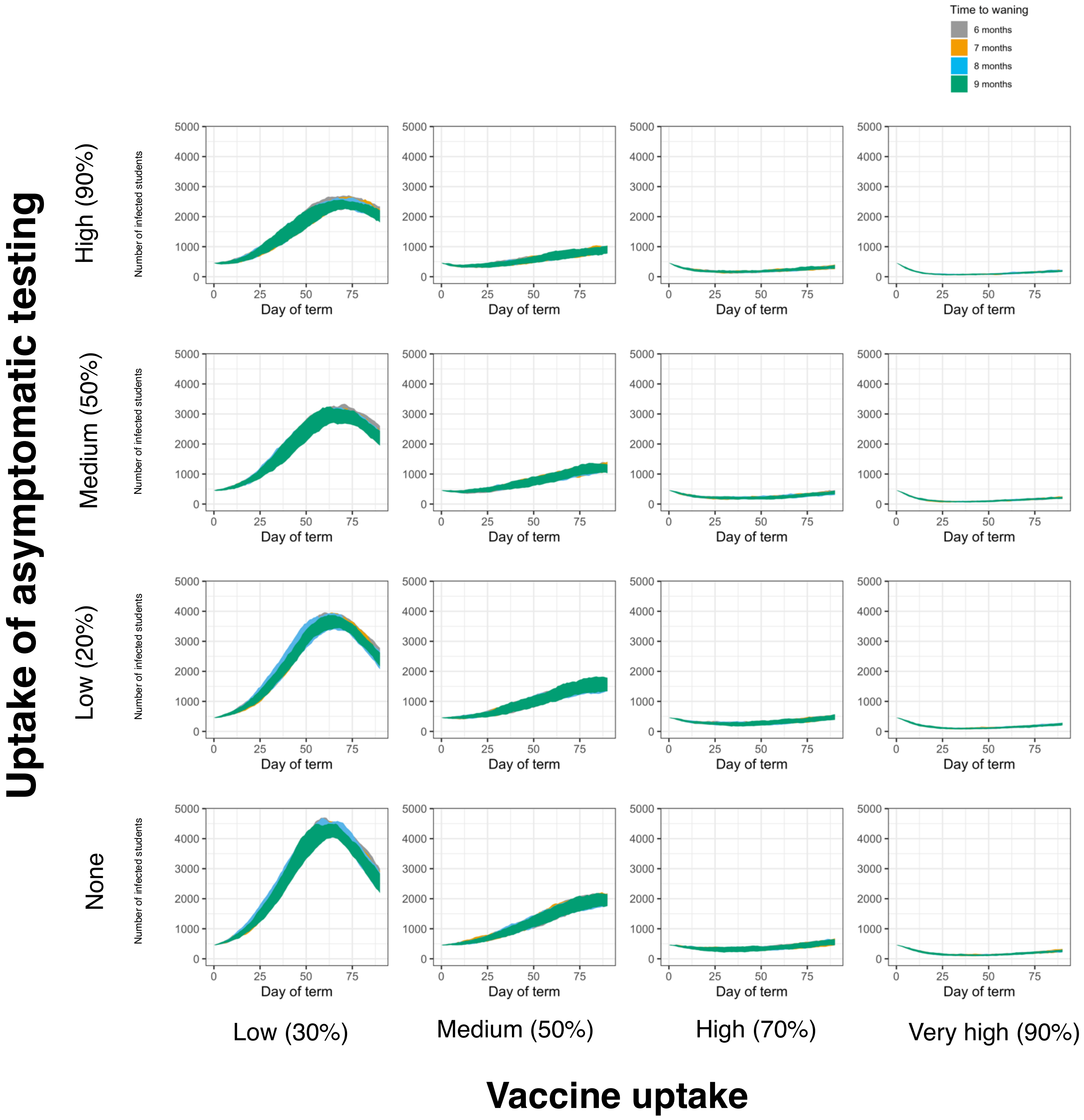


**Figure S14. Daily numbers of SARS-CoV-2 infections in a population of students (n=28116) during the autumn term of 2021 as predicted under different scenarios of waning immunity, vaccine uptake, asymptomatic testing and with 2% of students initially infected.** Each scenario is repeated 50 times and the range is given. Importation from the community was assumed to occur at a time-varying seasonal rate with maximum value of 1e-3.
